# Low Dimensional Chaotic Attractors in SARS-CoV-2’s Regional Epidemiological Data

**DOI:** 10.1101/2022.09.16.22280044

**Authors:** Carlos Pedro Gonçalves

## Abstract

**Background:** Recent studies applying chaos theory methods have found the existence of chaotic markers in SARS-CoV-2’s epidemiological data, evidence that has implications on the prediction, modeling and epidemiological analysis of the SARS-CoV-2/COVID-19 pandemic with implications for healthcare management.

**Aim and Methods:** We study the aggregate data for the new cases per million and the new deaths per million from COVID-19 in Africa, Asia, Europe, North and South America and Oceania, applying chaos theory’s empirical methods including embedding dimension estimation, Lyapunov spectra estimation, spectral analysis and state-of-the-art topological data analysis methods combining persistent homology, recurrence analysis and machine learning with the aim of characterizing the nature of the dynamics and its predictability.

**Results:** The results show that for all regions except Oceania there is evidence of low dimensional noisy chaotic attractors that are near the onset of chaos, with a recurrence structure that can be used by adaptive artificial intelligence solutions equipped with nearest neighbors’ machine learning modules to predict with a very high performance the future values of the two target series for each region. The persistent homology analysis uncovers a division into two groups, the first group comprised of Africa and Asia and the second of Europe, North and South America. For Oceania, we found evidence of the occurrence of a bifurcation which we characterize in detail applying a combination of machine learning and topological analysis methods, we find that the bifurcation in the region is related to the emergence of new variants.

## 1. Introduction

A virus contagion dynamics among human populations depends upon biological factors such as, among others, the virus characteristics, including incubation period, first time for symptoms to appear, the type of symptoms, the number of asymptomatic cases, morbidity, mortality, the mutation rates for the emergence of new variants, as well as the infected population and transmission vectors, on the other hand, there are also other factors that include a country’s population’s behavior and the measures put in place for healthcare response, quarantine, treatment, drugs, vaccination policies and, even, healthcare communication and public education on the virus and healthcare risks associated with it.

In this sense, since human societies respond to viruses and associated diseases with adaptive measures, there is a complex adaptive dynamics of human societies in response to virus contagion and outbreaks, response that, in turn, influences the viral propagation dynamics and the occurrence and pattern of outbreak waves, with possible complex attractors emerging from this dynamics. The nature and structure of these attractors may offer a basis for viral contagion and outbreak waves’ prediction and characterization using techniques coming from chaos theory, with possible value and insights for healthcare authorities, viral outbreak containment policies including possible quarantines, vaccination policies and healthcare resources planning.

Recent research into the Severe Acute Respiratory Syndrome Coronavirus 2 (SARS-CoV-2) has uncovered evidence of low dimensional attractors and markers of chaos in the pandemics’ dynamics for different countries and regions [1-4]. Taking into account this evidence, in the current work, we apply chaos theory’s main empirical methods including both chaos metrics such as embedding dimension and Lyapunov spectra estimation along with spectral analysis and expand on these empirical methods by employing state-of-the-art topological data analysis methods including: recurrence analysis metrics, persistent homology analysis, predictability metrics from nearest neighbors machine learning algorithms incorporated into adaptive artificial intelligences (A.I.), *k*-nearest neighbors graph analyses and ordinal partition graph analyses. These analyses allow us to provide a detailed characterization of the SARS-CoV-2’s dynamics per region. In terms of major findings, we uncovered the following key findings:

- Africa, Asia, Europe, North and South America show strong evidence of an emergent low dimensional noisy chaotic attractor that is near the onset of chaos with a black power law noise-like spectrum (color chaos) and a strongly predictable dynamics with a recurrence structure that can be used by forward looking adaptive A.I. solutions that can be employed to predict the future viral outbreaks;
- The topological data analysis techniques also uncover both common patterns to the regions as well as some level of diversity between regions, the persistent homology method, in particular, uncovers a division in two groups in terms of persistent of homology classes, the two groups are divided as follows: Africa and Asia on one group and Europe, North and South America on another group;
- Oceania shows evidence of a bifurcation with an increase in epidemiological risk that we characterize using machine learning tools and *k*-nearest neighbors graph analysis along with ordinal partition graph analysis.

The dataset that we use is the Our World in Data SARS-CoV-2 data, for the time period that goes from the earliest available data up to 2022-08-07, which was the last day available at the time of the analysis. The available dataset, thus covers the following periods for each region:

- Africa: 2020-02-13 to 2022-08-07;
- Asia: 2020-01-22 to 2022-08-07;
- Europe: 2020-01-23 to 2022-08-07;
- North America: 2020-01-22 to 2022-08-07;
- South America: 2020-02-22 to 2022-08-07;
- Oceania: 2020-01-25 to 2022-08-07.

The advantage of a regional division, while providing aggregate data, is that it produces a pattern that captures the general dynamics in the corresponding region in terms of general outbreak dynamics, while compensating for specific countries’ undercounting of cases, a major problem in the case of SARS-CoV-2 which limits the country-by-country analysis due to the need for test availability and the number of asymptomatic cases, a similar issue being raised for the number of deaths. In this way, the regional analysis provides for a way to mitigate this and for a better capturing of the general pandemic dynamics. A second point is that, while dealing with a pandemic, regional analysis allows one to identify commonalities and differences that may characterize different regions and explain co-outbreaks, that is outbreaks that occur almost simultaneously in different countries belonging to the same region.

The work is organized as follows:

- In section 2, we review the main concepts and methods used in the current work.
- In section 3, we apply the methods and tools reviewed in section 2 to the dataset.
- In section 4, we provide for a conclusion and final reflections of the work’s implications for epidemiological research into the SARS-CoV2’s pandemic and healthcare policies.

## 2. Main Concepts and Methods

### 2.1. Deterministic and Stochastic Chaos

Deterministic chaos can be defined as a bounded aperiodic dynamics with sensitive dependence upon initial conditions that exhibits a dynamical sequence that is random-looking with spectral signatures that are akin to a stochastic process [5-9], in this sense, deterministic chaos is often considered as a form of endogenous (pseudo)random dynamics in a deterministic system without any noise term, this type of chaos is closed in the sense that there is no external environment leading to external random noise.

In nature, especially when dealing with complex systems which are open and interact nonlinearly with an environment, we never have a fully closed deterministic process, noise is always present, this leads to stochastic chaos, which is an open chaos dynamics [7, 10, 11]. Within a purely mathematical setting, when it comes to random-like signals, for the purpose of systematization, there are three major dynamical processes [7, 10-12]: deterministic chaos, stochastic processes without a nonlinear chaotic component and stochastic chaos, which involves characteristics of the first two. An example of the last type is a system of nonlinear dynamical equations (in discrete time or in continuous time) that have a noise coupling.

The main problem with stochastic chaos is that noise is integrated into the dynamics and contributes to it [7,11]. The range of such a dynamics is very wide, one can have noise-induced chaos, an example of which is the destruction of periodic windows through noise, in a nonlinear dynamical system [11]. One can also have high dimensional or low dimensional attractors depending on the relation between the “external” stochastic component (the noise) and the “internal” nonlinear dynamical component [11, 12].

The type of noise coupling as well as the nature of noise also matters. Indeed, while we can have, for instance, a system of deterministic nonlinear dynamical equations that lead to a power spectrum equivalent to a white noise spectrum, we can also have deterministic chaotic systems that produce complex fractal and even multifractal signatures with power law decay in the power spectrum [11], if a such a deterministic chaotic dynamical system is coupled with a fractal or multifractal noise process, then the final fractal or multifractal signatures result from a complex interaction between the nonlinear dynamics and the complex noise process. Likewise, while high frequency white noise processes may be present, a deterministic chaotic dynamics with a power law decay in the power spectrum, which corresponds to 1/*f*^*β*^ chaos when coupled to a white noise process can lead to a 1/*f*^*β*^ noise signature with a high frequency breakdown, we see examples of this in financial market modeling in [11] and it corresponds to the concept of power law chaos.

Power law chaos, also called color chaos [11, 13], is relevant since it produces long range dependences, and, in the case of *β* > 2, it leads to the so-called black noise-like spectra, which are persistent processes that characterize different natural disasters [14]. In the epidemiological context, a black (stochastic) chaos dynamics, that is, (stochastic) chaos that produces signals with black noise spectra leads to persistent processes that can be characterized by long periods of low cases and then persistent large outbreak waves.

Empirically, stochastic chaos can be identified by the presence of positive maximum Lyapunov exponents in systems with dynamical noise, these signatures can show up either in the original signal or in a denoised signal. However, while the presence of these signatures in a noisy signal is indicative of a possible stochastic chaotic dynamical system, the absence of these signatures cannot also rule out the possibility of stochastic chaos.

This last issue is even more problematic for dimension estimations, indeed, we can have high dimensional chaos, a chaotic dynamics coupled to a high noise level, a coupling between a nonlinear dynamics and a complex stochastic process (for instance, fractal or multifractal noise), or even high dimensional chaos [11, 12]. By contrast, the emergence of a low dimensional attractor with positive maximum Lyapunov exponents in a noisy dynamics is indicative of a process of self-organization in a stochastic chaotic dynamics. In the empirical section we will find this to be the case for all regions except for Oceania, which exhibits a dynamical regime change corresponding to a bifurcation.

In complex adaptive dynamics such as an epidemic or a pandemic where human communities respond adaptively, an emergent chaotic process always falls into a stochastic chaos framework. The emergence of a low dimensional attractor, however, even in the presence of dynamical noise, implies the possibility of employing phase space reconstruction methods to predict the future dynamics of the series, especially in the case of black (stochastic) chaos, which leads to persistence patterns with recurrence structures and topological signatures that can be exploited by adaptive A.I. systems, exploiting topological features of the attractor in order to predict the future dynamics of relevant epidemiological targets.

In this case, the emergence of a low dimensional attractor, means that the interplay between the complex nonlinear dynamics has led to the emergence of a few robust small number of degrees of freedom that are supported by the system’s dynamics, and that may be linked to the adaptive dynamics themselves, leading to strong bottom-up and top-down connections [11, 12].

Thus, for instance, in the case of a virus spreading in a country, when the healthcare authorities calculate macroscopic variables, such as the number of new infected individuals, they can adjust their healthcare policies to these variables’ evolution, which leads to a change in that evolution itself, leading to a strong coevolutionary dynamics between human agents’ responses, viral evolution and the emergent patterns themselves, in this way the agents are adapting to the consequences of their own behavior, macroscopically monitored through statistics, and this adaptation in turn will change the outbreak dynamics.

The emergence of low dimensional attractors, in these contexts, with convergent stable dimension estimates implies that a macro-level dynamics with a small number of active degrees of freedom is being robustly sustained, which allows us to speak of a form of self-organization to a low dimensional stochastic/noisy chaotic attractor [12]. In a disease spread this is an important point, since it implies that a structural dynamics with a specific dimensionality (number of degrees of freedom) has emerged and is being sustained in the coevolutionary process. A type of dynamical profile that is researched upon, within the complexity sciences, by both synergetics and also in the networked chaos literature [11, 12].

We will see, in the case of the SARS-CoV-2, that, for the data sample available at the time of the analysis, all regions, except Oceania, are characterized by such low dimensional attractors’ emergence, however, the profile is not homogeneous with differences between regions as well as spectral and topological commonalities that may be linked to the virus characteristics. We also find evidence that is favorable to the attractors being close to a bifurcation point that corresponds to the *onset of chaos*, which opens up the possibility of occurrence of bifurcations, especially linked to new variants, in the case of Oceania we indeed find evidence of the occurrence of one such bifurcation, which leads to the need to employ topological analysis methods in conjunction with machine learning in order to be effectively addressed.

### 2.2. Main Methods

#### Phase Space Embedding

Phase space embedding is a method that can be used to uncover and reconstruct an attractor for a system’s dynamics from a time series. The main assumption is that the system’s dynamics may be described by an attractor in a *d*_*E*_ dimensional phase space, with *d*_*E*_ corresponding to the number of degrees of freedom. When dealing with complex adaptive systems, these dimensions correspond to emergent degrees of freedom resulting from coevolutionary dynamics leading to a geometrical structure that can be described by an attractor in a *d*_*E*_ dimensional phase space, the resulting attractor, which may contain a deterministic nonlinear component and dynamical noise, will depend upon the system in question [11, 12].

In general, the emergent attractor and corresponding phase space dimensionality are unknown, with researchers only having available a sample time series *x*_*t*_ which results from *T* sequential observations/measurements of the system in question, in this case, measurement noise and possible dynamical noise may be jointly present [12], however, assuming that there is an attractor in a *d*_*E*_ dimensional Euclidean phase space, the time series can be formally expressed as an observation function *x*_*t*_ = *g*(**y**_*t*_) where **y**_*t*_ is a point in the *d*_*E*_ dimensional Euclidean phase space. If **y**_*t*_ is in an attractor, which is a dynamical invariant, then, we know that the sequence **y**_*t*_ is a trajectory in the attractor, and *x*_*t*_ results from that trajectory [15].

If the equations of motion for **y**_*t*_ are known along with *g*, then a prediction of *x*(*t*) can be built by applying these equations, and the dynamical properties can also be addressed from the study of these equations and simulations of the system’s dynamics. However, when neither **y**(*t*) nor *g* are known this is no longer possible, in this case, delay embedding is a phase space reconstruction method working from the signal *x*(*t*) that is able to recover the main properties of the attractor, the method is based on Takens’ theorem [8, 15], using an appropriate time delay *τ* and embedding dimension *d*_*E*_ in Euclidean space, we can build the *d*_*E*_ dimensional tuples:

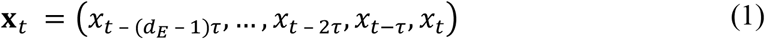

The resulting trajectory of the phase point **X**_*t*_ can recover, under certain conditions, the dynamics of the underlying unknown attractor [15]. The embedded trajectory can thus be used to study the main properties of the unknown attractor and as a feature space for time series’ prediction using machine learning techniques [8].

Methods for setting embedding parameters are aimed at obtaining an attractor reconstruction that captures the main properties of an underlying attractor, which means, in this case, having to choose values for the parameters *d*_*E*_ and *τ*.

The delay choice should be linked to the memory and characteristics of the dynamics. In epidemiological contexts, the choice of the delay should be defined in terms of the incubation period and quarantine window, setting the period to the first day after a recommended quarantine period allows for the embedding to account for quarantine effects.

In this case, we use, for *τ*, a 15 day period, the 15 day lag is the number of days between the start and the end of the recommended 14 day quarantine period. Indeed, the World Health Organization’s (WHO) recommended a 14 day quarantine period from a person’s last exposure, which leads to 15 days between the last exposure and an exit from quarantine [16]. While there were differences in the implementation of the quarantine period [17], the WHO recommended period became a policy standard and can be used as a baseline for delay setting, in this way, we are accounting in our delay choice for the quarantine effects, in an exploratory study we also found better results by working with the 15 period delay.

To choose the optimal embedding dimension, we use the lowest value for the false nearest neighbors [3, 18, 19]. The concept of false nearest neighbors is about topology and dimensionality, if the embedding dimension is too low with respect to the attractor’s dimensionality, then, one is obtaining a lower dimensional projection of a higher dimensional geometrical structure, in this case, points that are not neighbors will be projected onto a close neighborhood in the lower dimensional embedding, which will lead to problems, especially when dealing with topological data analysis, one of these problems is that what may seem to be noise-like signatures are associated not with noise but with the attempt to embed a higher dimensional object in a lower dimensional space.

When rising the number of phase space dimensions used in the phase space embedding, there is a reduction of false nearest neighbors, that is, phase points projected onto a close neighborhood of each other but that in a higher dimensional space embedding are found not to be neighbors at all, in this way, the number of false nearest neighbors tends to reduce as the number of embedding dimensions are risen, in a fully deterministic attractor, the false nearest neighbors will become zero when the dimension achieves the right number of dimensions needed to embed the attractor. When there is noise present, this percentage will not become zero but may be low, depending upon the strength of the deterministic component versus the noise level. In this way, the method is able to distinguish between low and high dimensional attractors and fully deterministic from stochastic dynamics.

In the case of a bifurcation, which happens for the Oceania’s series, we cannot, however, employ the false nearest neighbors method, because we are not dealing with a stable attractor, in this case, we need to employ a different methodology which will use machine learning and the topological data analysis methods addressed further on, this methodology is presented and discussed in section 3 when we address the Oceania’s case.

#### Lyapunov Exponents Estimation

A positive largest Lyapunov exponent, is an indicator of sensitive dependence on initial conditions and a marker of chaos. In the present work, we use Eckmann and *et al*.’s method for Lyapunov spectrum estimation [20], which estimates the spectrum of Lyapunov exponents for a multidimensional attractor. For the complete and detailed description of the algorithm we refer to the original reference [20].

The method allows one to set a matrix dimension *d*_*M*_ and study the behavior of the exponents for increasing embedding dimensions *d*_*E*_ ≥ *d*_*M*_, we can set the dimension *d*_*M*_ equal to our “suspected” dimensionality of the attractor. An advantage of Eckman *et al*.’s method is that it is able to extract a Lyapunov spectrum for the set matrix dimension, with one exponent for each dimension, the identification of positive exponents is evidence favorable to chaos. In this case, if we find a convergence of the Lyapunov spectrum as the embedding dimension increases and some of the exponents converge to positive values, then, this is evidence favorable to chaos [20].

#### Topological Data Analysis Methods

Topological data analysis methods are perhaps the most sophisticated and robust methods in nonlinear time series analysis, especially for short time series. These methods allow one to better characterize the dynamics and distinguish between periodic, quasiperiodic, chaotic and random dynamics as well as address bifurcations.

The main structure is the distance matrix *S* obtained for an embedding with entries given by the distance between any two points of an embedded trajectory in a *d*_*E*_ dimensional Euclidean space. This matrix has entries given by:

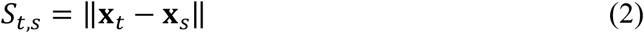

Different distance metrics can be used, the most common being the Euclidean metric [8, 21], which takes advantage of the Euclidean metric space topology and has some advantages for the methods we will implement here. The matrix *S* provides for information on the recurrence structure of the dynamics, becoming a relevant source of topological information on the attractor’s structure. Taking advantage of the Euclidean metric topology of the embedded trajectory, the distance matrix can be used to produce a neighborhood radius-dependent binary recurrence matrix, this is the *ε*-recurrence matrix *B*_*ε*_ with entries:

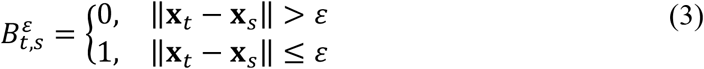

This is a square symmetric matrix that registers the value 1 when two phase points do not differ by more than *ε*, and 0 otherwise. The use of the closed neighborhood structure, in the recurrence analysis, has the advantage of allowing us to identify fully periodic dynamics so that, in the case of a periodic dynamics, if the radius is equal to zero, all diagonals parallel to the main diagonal that differ from each other by the period in question will have the value of 1 in each matrix entry, otherwise the value will be 0 [21]. In the nonperiodic case, we never get these evenly spaced full diagonals of radius of 0.

We will perform several analyses on the distance matrix, the first is the calculation of two metrics [21]: the recurrence strength and the conditional 100% recurrence probability. The recurrence strength is the sum of the number of points that fall within a distance no greater than the radius, in each diagonal below the main diagonal, divided by the total number of diagonals below the main diagonal with recurrence, this measure evaluates how strong on average the recurrence is. The conditional 100% recurrence probability is the probability that a randomly chosen diagonal line with recurrence has 100% recurrence, for the radius chosen. If all lines with recurrence had 100% recurrence, for the radius chosen, then this number would be equal to 1, the lower this metric is, that is, the closer to zero it is, the more interrupted the diagonals are, which occurs for stochastic systems and also for deterministic chaotic dynamics [21].

Chaotic dynamics, depending upon the strength of the exponential divergence characterized by the largest Lyapunov exponent and the dimensionality of the attractor, usually displays a higher recurrence strength and, with rising radius, it can, in general, exhibit a higher conditional 100% recurrence probability than stochastic processes. We calculate the above two metrics for different radii in order to characterize the topological properties of the attractor in terms of neighborhood structure, using radii that are multiples of the series’ standard deviation.

The second type of analysis that we perform is based on persistent homology. In this case, the distance matrix *S* for the phase space reconstructed trajectory sets up a family of transition matrices corresponding to the *ε* recurrence matrices *B*_*ε*_, the resulting transition graphs are, thus, based on the topology induced by the Euclidean metric (if the Euclidean metric is used to construct the distance matrix *S*). Now, relevant topological structures can be extracted by working with the distance matrix to build a parametrized family of simplicial complexes *C*_*ε*_ obtained from the sequence of *ε* recurrence matrices *B*_*ε*_, with *ε* ≥ 0, with an edge connecting two vertices, corresponding to phase points **X**_*t*_ and **X**_*s*_, being included when 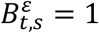, therefore, at any given radius, different phase points at different times (even distant times) may be close in value, that is, they may be neighbors in the attractor, and, thus, linked by an edge in the simplicial complex. For increasing radii *ε*′ ≥ *ε* the parametrized family leads to a Vietoris-Rips filtration: *C*_*ε*_ ⊆ *C*_*ε*′_ ⊆ ⋯ [22, 23].

Persistent homology, as a topological data analysis tool, when calculated on a reconstructed attractor’s full distance matrix *S*, which includes all the Euclidean distances between the points, allows one to find relevant topological features in the reconstructed attractor and analyze how the homology changes over the filtration. Homology theory is a branch of algebraic topology that studies the connectivity properties of topological spaces [23]. In a simplicial complex, we can look at different homology dimensions and corresponding homology class, a 0 homology class (H0) corresponds to components connected by a path (which leads to a zero dimensional boundary), a 1 homology class (H1) corresponds to a loop, voids which are simplexes with faces but no interior correspond to a 2 homology class (H2), and so on [23].

Persistent homology can be used to count the number of structures in each simplicial complex in the filtration for each homology dimension, including the birth and death of homology classes as the radius is increased. The homology classes’ birth and death can be calculated in the following way, given a filtration of simplicial complexes 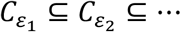, we get a sequence of maps for the homology dimension *s*, 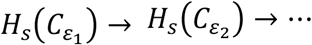, a homology class is born at *n* if it is in 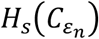 but not in the image of the map 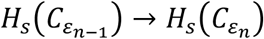 and dies at *m* if it is in 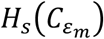 but not in 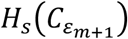 [23].

Persistent structures have “long lives” in the filtration and correspond to large scale structures in an attractor, structures with lower persistence usually appear in noisy and chaotic systems. A persistence diagram *D* can be calculated for different homology dimensions with each point for each dimension giving the lifetime for a structure, in this case, we define *D*_*s*_ as the persistence sub-diagram for the homology dimension *s*, each point in the sub-diagram corresponds to an ordered pair of birth and death times in the filtration.

The lifetime or persistence of a class at dimension *s* corresponds to the difference between the death and birth times, thus, given a dimension *s* and the ordered pairs (*n*_*B*_, *n*_*D*_) ∈ *D*_*s*_, where *n*_*B*_ is the “filtration birth time” and *n*_*D*_ is the “filtration death time”, with the death happening after birth, we can calculate the persistence as

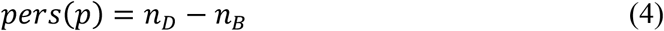

Structures that live through the full filtration have *n*_*D*_ = ∞ and, therefore, *pers*(*p*) = ∞. Now, to better characterize the attractor’s topology we calculate the following metrics for each homology dimension (that is, for each sub-diagram *D*_*s*_):

- The number of classes with *pers*(*p*) < ∞: this metric allows us to identify which dimension is predominant in terms of number of homology classes with lifetimes shorter than ∞;
- The number of classes with *pers*(*p*) = ∞: this metric allows us to identify the homology dimensions that have structures that persist throughout the whole filtration, constituting large scale structures;
- The maximum persistence: this metric allows us to characterize each homology dimension in terms of the maximum persistence, which allows us to characterize which dimensions have the most persistent structures;
- The mean persistence: this metric allows us to characterize each homology dimension in terms of its mean persistence.

With these metrics calculated for the different sub-diagrams we get a picture of the attractor’s structure at multiple dimensions and the dominant features.

Now, as another topological data analysis method, in order to evaluate the degree to which the recurrence structure in the Euclidean distance matrix *S* supports a prediction of future values of the target series, we can use a nearest neighbor machine learning method to build an adaptive A.I. system to predict the series *k* steps ahead using the Euclidean radius to predict the next step, this is nearest neighbors-based regression that uses a radius to predict the next steps in a series.

In the current work, we use Python’s machine learning module scikit-learn’s radius neighbors regressor for this task. We stress that the main objective of this application is not to test different machine learning methods in the prediction of the new cases per million and the new deaths per million series, but, instead, our aim is to evaluate the degree to which topological information associated with the Euclidean recurrence structure contained in the matrix *S* can be used to predict the target epidemiological series, this explains the choice of machine learning architecture which must be one that uses the Euclidean radius to predict the target series, therefore, the employment of the machine learning here is as an added topological data analysis method.

Given a phase space embedding, we can apply a radius neighbors-based regression using the embedding of a past trajectory as a dynamical feature space, and perform the regression over a learning sliding window of a reconstructed orbit {**X**_*t*−*w*_, …, **X**_*t*−1_} and the sequence of observed signals {*x*_*t*−*w*+*k*_, …, *x*_*t*+*k*_}, for *k* = 1,2, …, used as training data so that the A.I. learns window-dependent prediction function:

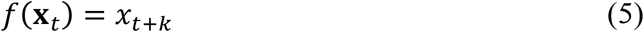

We can then use the learned function to predict the target series *k* steps ahead by applying the function *f*(**X**_*t*_), the sliding window is then moved one step to the right and the learning algorithm is run again. This is a basic prospective prediction algorithm where the A.I. system behaves as an adaptive agent that relearns the patterns and adapts to changes in the series, adapting to the recurrence structure. Sliding window prospective models have been successfully applied in epidemiological prediction, a noticeable model was the testing of a sliding window prospective prediction model used to predict the H5N1 dynamics by Kane *et al*. [24].

This adaptive agent prospective method is more effective than the fixed training data method for small datasets and for dealing with complex systems’ dynamics that exhibit bifurcations, attractors that have long periods, attractors with changing recurrence epochs, including alternation between laminar periods and turbulent periods, as well as regime switching. In this way, the adaptive agent approach allows the agent to capture epoch-specific recurrences in sliding windows (sliding recurrence epochs) and also adapt to changing dynamical patterns.

Again, since our goal is to evaluate the degree to which the recurrence structure in the attractor contains information on the future dynamics, which is a hallmark of the presence of a deterministic structure [8], we quantify the prospective adaptive agent’s performance using as main metrics the coefficient of determination, explained variance, root mean squared error divided by the data amplitude and the linear correlation between the observed and predicted values, in order to evaluate the degree to which the recurrence structure contains information on the future series’ dynamics.

Now, besides the distance matrix *S*, there are two other structures for topological analysis that can be calculated from the reconstructed attractor, the first is the *k*-nearest neighbors graph **N**, the second is a symbolic dynamics tool called the ordinal partition graph **O** [23] which is also used to calculate the permutation entropy [23, 25].

In the first case, we have an undirected graph with the vertices corresponding to each phase point and the edges corresponding to the *k* nearest neighbors [23]. To select the number of *k* neighbors to analyze, we use another machine learning method to build an adaptive agent, with a similar approach as described above, but instead of a radius learner we use a *k*-nearest neighbors’ learner for a single step prediction, by evaluating its performance on different *k* values we choose the value of *k* that leads to the highest predictability, since that is the value which contains the highest information for predicting the target series. In this case, we choose the value of *k* that leads to the highest coefficient of determination and then analyze the resulting *k*-nearest neighbors’ graph, calculating the degree entropy and the Kolmogorov-Sinai (K-S) entropy for the graph [26].

Given a general graph *G* comprised of *n* nodes and the degree associated with each node, we can extract a frequency distribution over the set of different degree values *S*, leading to the relative frequencies *p*_*s*_ associated with each degree value *s* ∈ *S*, from this distribution we can calculate the degree entropy as:

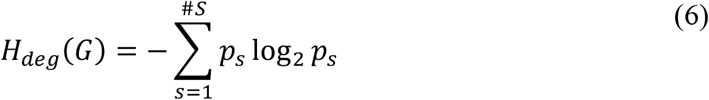

In the special case of a graph where the relative frequencies associated with each degree coincide *p*_*i*_ = 1/#*S*, we get the maximum entropy, with *H*(*G*) = log_2_ #*S*, in the case of a graph where each node has the same degree we get *H*(*G*) = 0. To obtain a value between 0 and 1 we can work with the relative entropy:

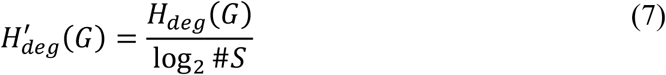

The closer to 1 this last degree entropy measure is, the closer the graph is to a maximum degree entropy. From now on, when we refer to the degree entropy, we mean this last definition.

The K-S entropy provides, on the other hand, for an important synthesis for a dynamical system, since it is an information measure on the sequence of nodes for a Markov process on a network, in this case, for the *k* nearest neighbors graph it provides an information measure for a Markov process with a transition matrix extracted from the graph, likewise, for the ordinal partition graph it provides for an information measure on a Markov process with a transition matrix extracted from the transition graph between the different permutations. For an unweighted graph, which is our case, this entropy coincides with the logarithm of the dominant eigenvalue of the transition matrix *μ*_+_, expressing it in bits leads to the following information measure [26]:

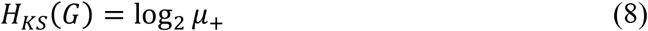

Now, besides the *k*-nearest neighbors graph, there is a final topological analysis method that is relevant, which is the, already mentioned, ordinal partition graph which is built from the permutations 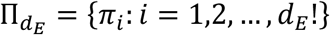 of the dimensions’ set {1,2, …, *d*_*E*_}, in this case, a permutation map 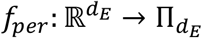 is defined such that 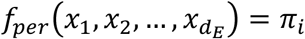 with π_*i*_ satisfying the condition that 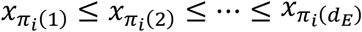, that is, the permutation produces a nondecreasing reordering of the values in the *d*_*E*_ tuple of real numbers, now, evaluating the permutation map for each phase point **X**_*t*_, we get a sequence of permutations *f*_*per*_(**X**_*t*_), from this sequence we can build the ordinal partition graph **O**, where two permutations are linked if there is a transition between them.

The permutation entropy can be calculated from the probability distribution over the permutations in the ordinal partition graph **O** [23,25]:

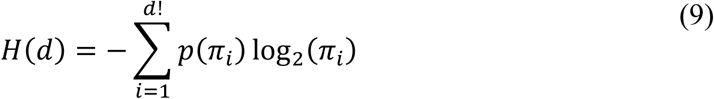

For the ordinal partition graph **O**, we also calculate here the degree entropy and the K-S entropy.

## 3. Results

### 3.1. Analysis of Series with Attractors

The analysis we perform, as discussed in the introduction, is focused on the regional division in terms of Continents with an additional subdivision between North and South America. It turns out that a regional division, while providing aggregate data, produces a pattern that captures the general dynamics in the corresponding region in terms of general outbreak dynamics while compensating for specific countries’ undercounting of cases, a major problem for SARS-CoV-2, which limits the country-by-country analysis, and may be linked to test availability and the number of asymptomatic cases.

In an exploratory study, we found dynamical markers of undercounting, especially in Africa in both new cases and, in particular, in new deaths, leading some countries to exhibit a series with long zero new identified cases, and zero new deaths interrupted by a few jumps as well as ‘patchy’ series, while the overall aggregate pattern by region agrees for those countries that were able to implement a higher testing and were able to monitor the virus more closely. In this way, we work with the aggregate pattern by region, with similar methodology being possible to implement in those countries where the testing frequency allows for sufficient fluctuations to apply the analysis, providing for a basis for comparison between regions that otherwise would not be possible.

In figure 1, we show the time series plots for the number of new cases per million for each region, obtained from the Our World in Data website. As can be seen from the plots, Africa stands out with a fast pattern of large successive waves this is the multiple wave pattern, while Asia, Europe, North and South America have a series of waves and then a few very large waves, this corresponds to a rogue wave-like pattern, where we have a series of smaller waves and then we get a very large wave, in Oceania, on the other hand, we have a dynamical regime change, there is a region with smaller fluctuations and then a rogue wave formed, after which the pattern becomes more turbulent with a higher number of new cases per million.

**Figure 1:**
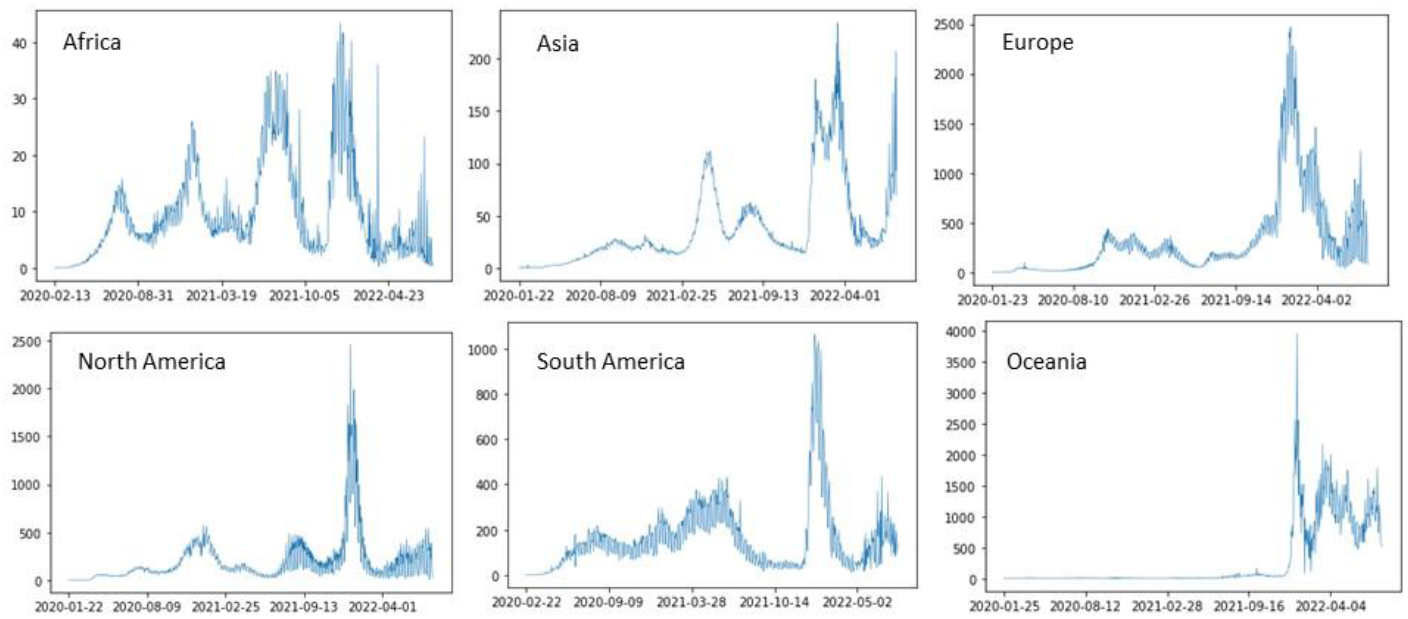
Number of new cases per million for each region in the database from the first observation to 2022-08-07.

The dynamical regime change in Oceania can also be seen in the new deaths per million plot, with a rise in turbulence and deaths after the regime change. This means that Oceania will require a different analysis process that will rely on the characterization of the regime change and a research into that change.

For the remaining five regions, which do not exhibit evidence of a dynamical regime change, we find evidence of a low dimensional chaotic attractor in both the number of new cases per million and the number of new deaths per million, as shown in tables 1 and 2. For the new cases per million series, the number of degrees of freedom for the emergent attractor is three with an associated small but non-null percentage of false nearest neighbors, this may be indicative of the presence of noise and consistent with a stochastic chaos process, the largest Lyapunov exponent, in each case, is positive and near 0.002.

**Table 1:** Main chaotic time series metrics for the five regions and new cases per million.

|  | **Embedding**<br>**Dimension** | **% FNN** | **$L_1$** |
| --- | --- | --- | --- |
| **Africa** | 3 | 6.364749 | 0.002112 |
| **Asia** | 3 | 4.423749 | 0.002160 |
| **Europe** | 3 | 6.162791 | 0.002674 |
| **North America** | 3 | 8.591885 | 0.002600 |
| **South America** | 3 | 5.816832 | 0.001877 |

**Table 2:** Main chaotic time series metrics for the five regions and new deaths per million.

|  | **Embedding**<br>**Dimension** | **% FNN** | **$L_1$** |
| --- | --- | --- | --- |
| **Africa** | 5 | 0.0 | 0.001575 |
| **Asia** | 3 | 3.716609 | 0.001939 |
| **Europe** | 4 | 0.704225 | 0.001490 |
| **North America** | 5 | 0.245098 | 0.001214 |
| **South America** | 4 | 1.472393 | 0.001314 |

For the new deaths per million, we also have evidence favorable to a low dimensional chaotic attractor for each of the five regions, however, the embedding dimensions differ between regions, indeed, while Asia is still characterized by a three-dimensional embedding, Europe and South America lead to a four-dimensional embedding, while for Africa and North America we get a five dimensional embedding. The percentage of false nearest neighbors in each case is very small, below 1%, with the exception of Asia which has an around 3.7% value and South America which has an around 1.5% value. Noticeably, Africa has a 0% estimate of false nearest neighbors. These results point to a lower noise level in the reconstructed attractors, when compared to the new cases per million, despite, in general, the higher dimensionality of the resulting attractors as estimated by the lowest percentage of false nearest neighbors. The largest Lyapunov exponents are all positive but smaller than those of the new cases per million.

The Lyapunov spectrum, as shown in figure 3, exhibits a good convergence with a dominant positive exponent in each region. In the new cases per million series, the Lyapunov spectrum converges to a positive exponent and two negative exponents, leading to a negative sum in the spectrum, which is consistent with dissipative chaos and the existence of an attractor in phase space in the number of new cases per million (figure 3 (left)).

**Figure 2:**
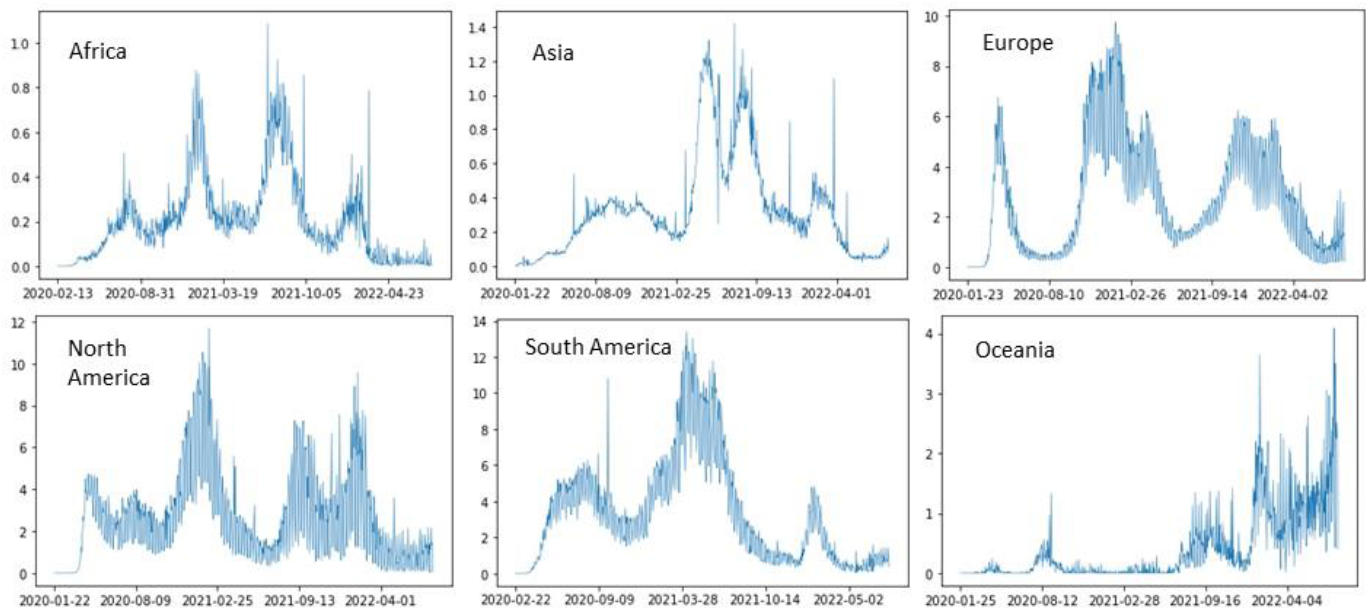
Number of new deaths per million for each region in the database from the first observation to 2022-08-07.

**Figure 3:**
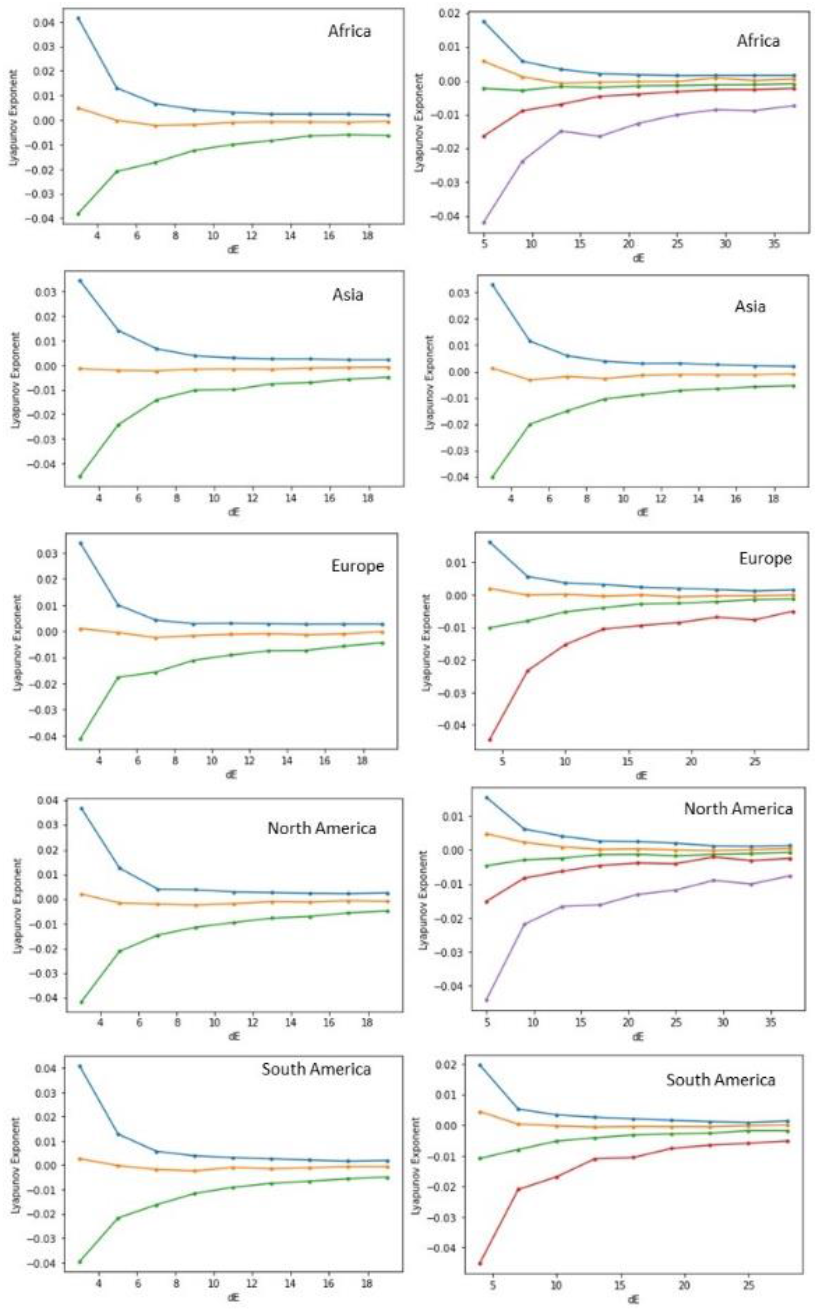
Lyapunov spectrum for the different regions’ embedded number of new cases per million series (left) and the number of new deaths per million (right), using matrix dimension equal to the dimension estimated by the false nearest neighbors with the spectrum calculated for increasing embeddings.

In the case of the number of new deaths per million’s Lyapunov spectra (figure 3 (right)), for Africa there is only one positive exponent, with the second largest exponent converging to a negative value but very near zero, a similar profile is obtained for Europe, Asia and South America, in the North America case, the second exponent converges to a positive value, however, it is also very near zero (0.000333). In each case the spectrum sum is negative which is, again, consistent with dissipative chaos and a chaotic attractor.

Applying spectral analysis to each series and region we find evidence of a black power law noise-like spectrum, in the majority of regions, in figure 4 we show the power spectra for Africa in log-log scale with the estimated slope.

**Figure 4:**
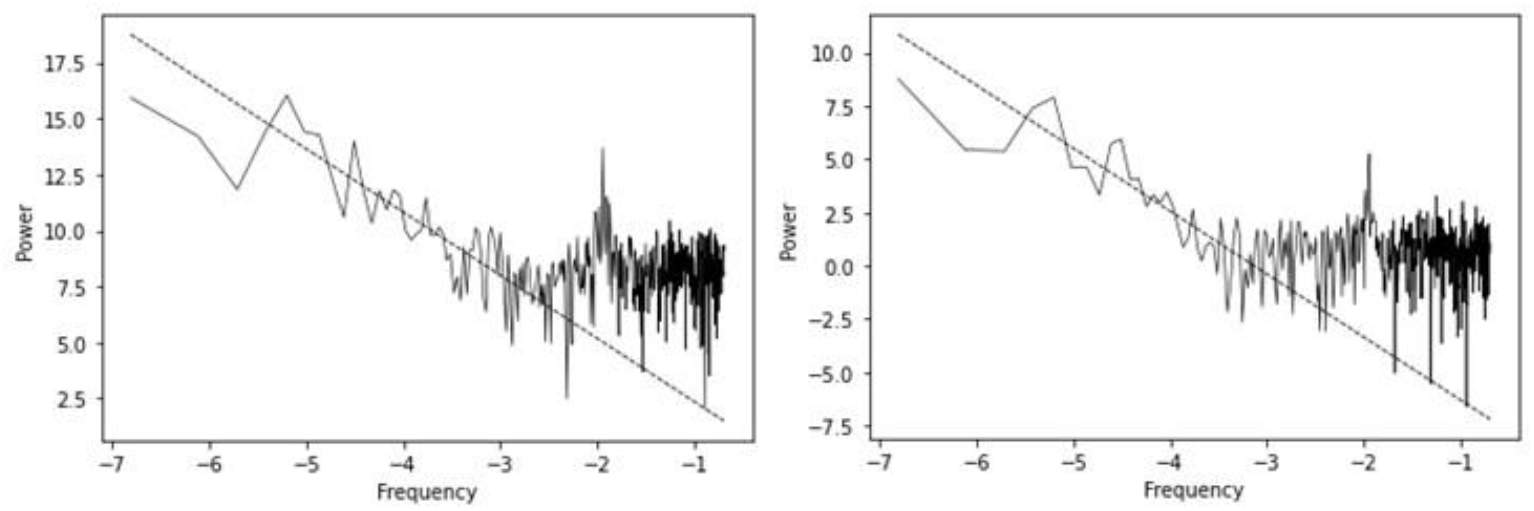
Power spectrum for Africa plotted in log-log scale with fitted line in the power law decaying region for the new cases per million series (left) and the new deaths per million series (right).

The estimated slope for the new cases per million in the power law decay region is -2.820713, with an associated *R*^2^ of 0.689679 and a *p-value* of 2.454008e-15, this means that we have 1/*f*^*β*^ spectrum with *β* approximately equal to 2.820713 > 2, which corresponds to a black power law noise-like spectrum. At the high frequency region we get a white noise spectrum with the exception of a rise in the spectrum and a peak that is consistent with a periodicity present at the high frequency region.

The estimated slope for the new deaths per million in the power law decay region is -2.953122, with an associated *R*^2^ of 0.674452 and a *p-value* of 2.731255e-12, again we have a black noise-like spectrum. Contrasting with the new cases per million, the new deaths per million, while exhibiting a similar peak at the high frequency region, also show a wider white noise region than the new cases per million.

The black noise power law decay implies a strong persistence which is linked to long memory and the formation of persistent large outbreak waves visible in both the number of new cases and deaths. Since the evidence is favorable to a low dimensional noisy chaotic dynamics, the black noise-like spectrum may be linked to the chaotic dynamics itself, in this case, to a form of color chaos [11, 13] that can be called black chaos since it has a black (power law) noise-like spectrum. Power law chaos occurs for attractors such as the Lorenz attractor as well as for nonlinear chaotic maps and characterizes the color chaos dynamics [11,13], with black chaos being a form of color chaos that exhibits a black noise-like spectrum.

The peak at the high frequency region may have different possible explanations, one possible explanation would be an artifact produced by a testing periodicity, but this hypothesis is not supported by the evidence given that we have a similar peak in the new deaths per million.

Considering the evidence, we have a low dimensional chaotic attractor for both series (new cases and deaths) comprised of a long-range dynamics, which is supported by the low false nearest neighbors’ values leading to a low embedding dimension, a positive albeit small largest Lyapunov exponent, and a black noise-like spectrum associated with the large outbreak waves and strong persistence that allows us to classify the dynamics as black chaos.

Now, the white noise spectrum at the high frequency region, after the power law decay, is either a feature of the attractor or a possible feature of the interplay between the black chaos and a white dynamical noise with a high frequency dominance, in which case, the peak may be a high frequency marker coming from the long memory chaotic process with power law spectrum (color chaos) leaving a dynamical marker in the form of a signal with a periodicity at the high frequency region, a signal that results from the interplay between the white dynamical noise and the underlying chaotic dynamics, this peak occurring thus as a form of noise-induced order in what is an underlying stochastic chaos dynamics.

Another possible explanation for the peak is that the chaotic attractor is close to a bifurcation point between a periodic orbit and a chaotic orbit. This possible explanation is supported by one piece of evidence, specifically, the low value of the largest Lyapunov exponents, which coupled with these spectral signatures support the possibility of the dynamics being near a bifurcation point corresponding to the *onset of chaos*, being close to a cycle, that is, the dynamics may be close to a bifurcation point between a periodic window and a chaotic dynamics, such peaks in the power spectra can indeed happen for chaotic dynamics close to the *onset of chaos*. Such a proximity is a hypothesis that fits the data well, furthermore, in stochastic chaos, the stochastic component may induce a transition from what would be a periodic orbit to a chaotic orbit, in such a way that we have a form of noise-induced chaos but with the resulting dynamics being close to the *onset of chaos*.

In chaos theory, the *onset of chaos*, as stated, corresponds to a bifurcation point where there is a transition from a periodic or quasiperiodic dynamics to a chaotic dynamics, near the *onset of chaos* we can find dynamics that, while being characterized by chaotic attractors, intermix long range periodic or quasiperiodic signatures in the recurrence structure for a sufficiently high value of the radius, this is so because near the bifurcation point, the chaotic dynamics has a low Lyapunov exponent and can exhibit recurrences associated with close proximity to periodic or even quasiperiodic orbits, especially if the dynamics is very close to the *onset of chaos* which is usually preceded by a regular structure such as, for instance, a periodic window.

Chaotic attractors near the *onset of chaos* tend to have low values for the maximum Lyapunov exponents and may recurrently visit a cycle or quasiperiodic orbit which becomes like a “ghost trail”, this sometimes shows up in the recurrence analysis in the form of long evenly (periodic) or unevenly spaced diagonals with 100% recurrence that only show up for a sufficiently high radius, diagonals that are intermixed with broken diagonals and isolated points which are characteristic of chaotic dynamics [21].

Considering, now, the case of Asia, in figure 5 is shown the power spectrum for the number of new cases per million and for the number of new deaths per million. We find again evidence of power law scaling with 1/*f*^*β*^ noise signatures with *β* > 2, again favorable to the black chaos hypothesis. For the new cases per million, this power law scaling is dominant with the breakdown to a white noise spectrum being highly reduced and confined to the high frequency region, also we still get a peak indicating a close proximity to a cycle.

**Figure 5:**
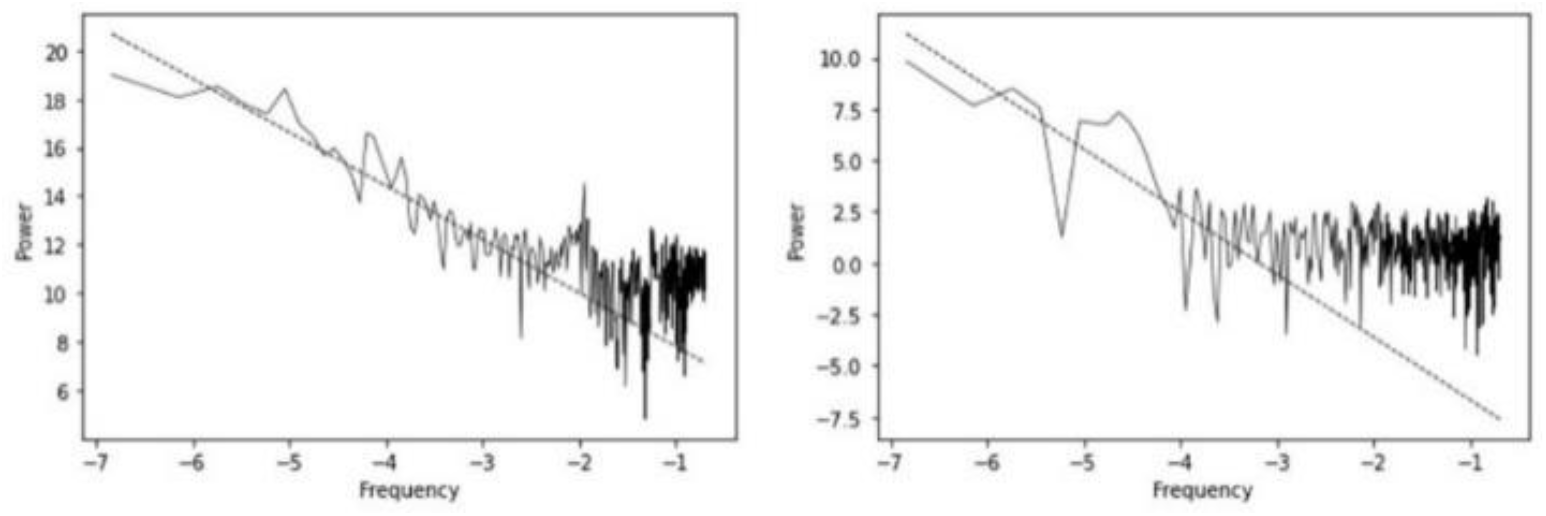
Power spectrum for Asia plotted in log-log scale with fitted line in the power law decaying region for the new cases per million series (left) and the new deaths per million series (right).

The estimated exponent for the new cases per million is not as high as that of Africa, being closer to 2, which also means that the dynamics is slightly less persistent for Asia than for Africa, indeed, the estimated exponent is 2.209483 (obtained from the estimated slope in the log-log plot), with an *R*^2^ of 0.850672 and a *p-value* of 1.265705e-25.

The new deaths per million series also shows a power law scaling, however the scaling corresponds to a much lower part of the spectrum, with the dominant frequency range being characterized by a white noise spectrum, which is consistent with the possibility that we are dealing with stochastic chaos where the dynamical noise process is characterized by high frequency white noise, a point already raised for Africa. The estimated exponent for the power law section is 3.058617, with an associated *R*^2^ of 0.600340 and a *p-value* of 5.009210e-07.

In the Asian case, there is a greater difference between the new deaths per million series and the number of new cases per million series, indeed, while in Africa we get a similar power spectrum profile, for Asia the power spectrum profile differs significantly, which contrasts with the dimensionality change, indeed, in Africa we go from a three dimensional to a five dimensional embedding, while in Asia the estimated dimensionality is the same.

Considering now the case of Europe, as shown in figure 6, we find a power law scaling for both the number of new cases per million and the new deaths per million, with a similar overall profile in what regards both the scaling and the high frequency region. The power law scaling, in both cases is the dominant part of the spectrum and in both cases corresponds to a black noise-like spectrum. For the new cases per million the estimated exponent is 2.879192, with an *R*^2^ of 0.865065 and a *p-value* of 6.643054e-27, for the new deaths per million we get a similar power law decay of 2.812378, with an *R*^2^ of 0.735955 and a *p-value* of 2.046621e-18, therefore, we have a similar decay profile. Furthermore, in the high frequency region, we do not have a white noise spectrum but rather two dominant peaks in the new cases per million and three peaks in the number of new deaths per million, which again points to the presence of strong periodicities at the high frequency level, both in the new cases per million and the new deaths per million.

**Figure 6:**
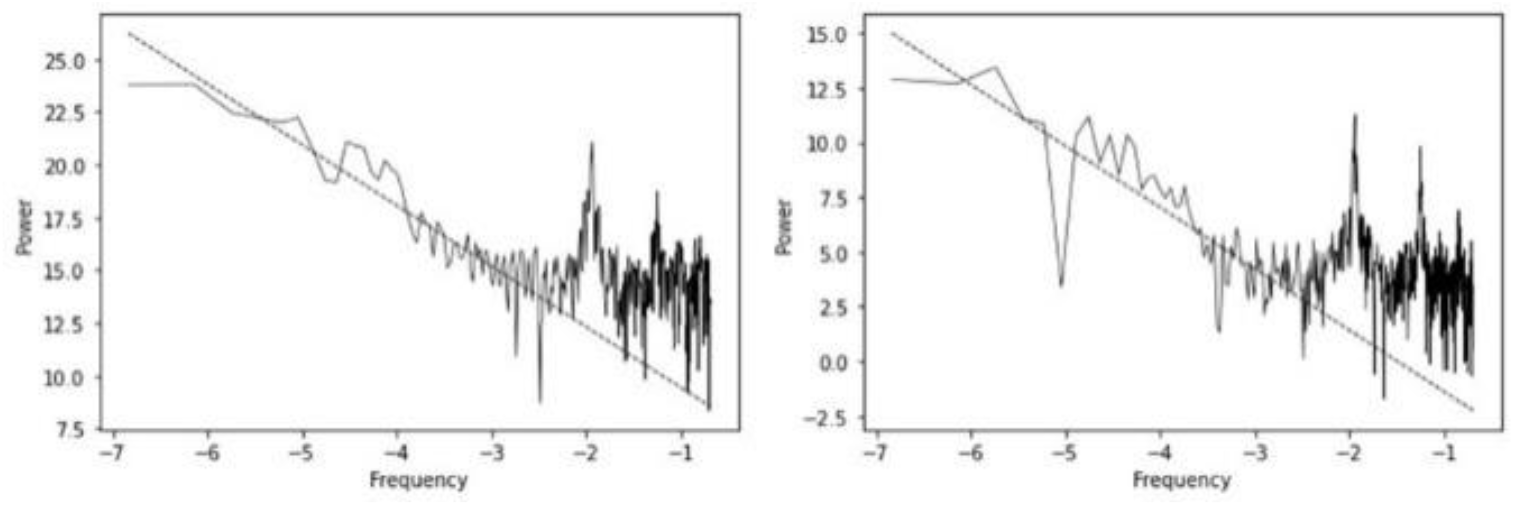
Power spectrum for Europe plotted in log-log scale with fitted line in the power law decaying region for the new cases per million series (left) and the new deaths per million (right).

Given the above results, we are led to the same hypotheses as those raised for Africa, one being that the peaks indicate the occurrence of some form of noise-induced order in a chaotic dynamics, the second being that the dynamics is close to a bifurcation point with the periodic signatures arising from the close proximity to a periodic window, leaving a marker at the high frequency part of the spectrum. Given the low value of the largest Lyapunov exponent and the fact that spectral peaks linked to periodicities can happen for a chaotic dynamics near a periodic window (dynamics near the *onset of chaos*) the last hypothesis, again, fits well with the data.

Considering, now, North and South America, we find different profiles than those obtained for the previous regions. In North America, the decay is faster than the power law, the high frequency spectrum is also not white noise but has three peaks in both the new cases per million and the new deaths per million. In this way, we find that there is some persistence, in the low to mid frequency but it is not power law scaling, and at the high frequency we have the evidence of multiple periodicities. Again, this is characteristic of a chaotic dynamics near a bifurcation point, the presence of these peaks in both the new cases per million and the new deaths per million reinforces the hypothesis of a dynamics near the *onset of chaos*.

As shown in figure 8, South America also has a fast decay for the new cases per million, but it has a power law decay in the deaths, the estimated exponent is 3.135302, with an *R*^2^ of 0.8566170 and a *p-value* of 2.492670e-13. Despite the different low frequency decay patterns, in the high frequency range we get two peaks, which is again consistent with the hypothesis of a close to the *onset of chaos* dynamics.

**Figure 7:**
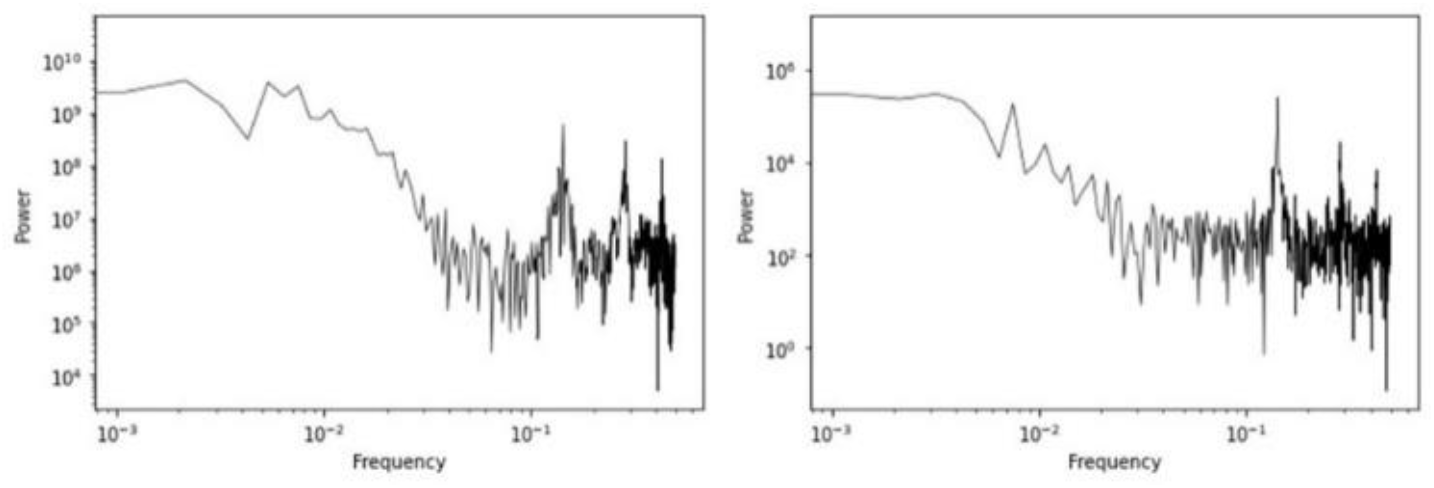
Power spectrum for North America plotted in log-log scale for the new cases per million series (left) and the new deaths per million (right).

**Figure 8:**
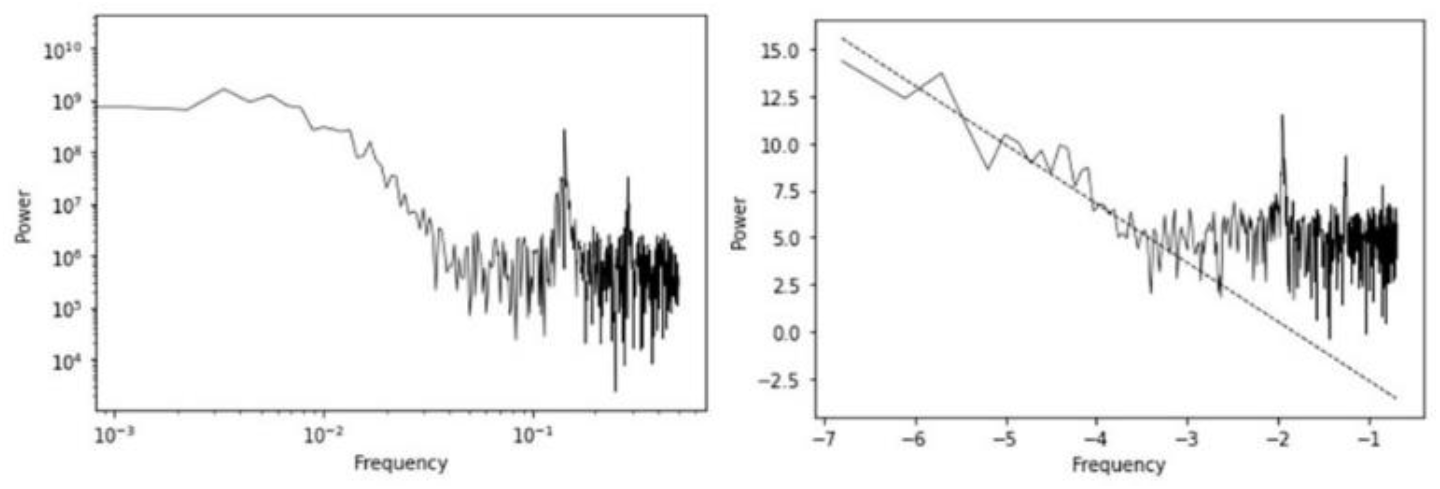
Power spectrum for South America plotted in log-log scale with fitted line in the power law decaying region for the new cases per million series (left) and the new deaths per million (right) with the fitted line in the power law decaying region.

Taking into account these results, we now turn to the topological analysis. In figure 9, we show the recurrence strength calculated for different radii with increasing radius in units of standard deviation (s.d.) for the number of new cases per million’s reconstructed attractors.

**Fig 9:**
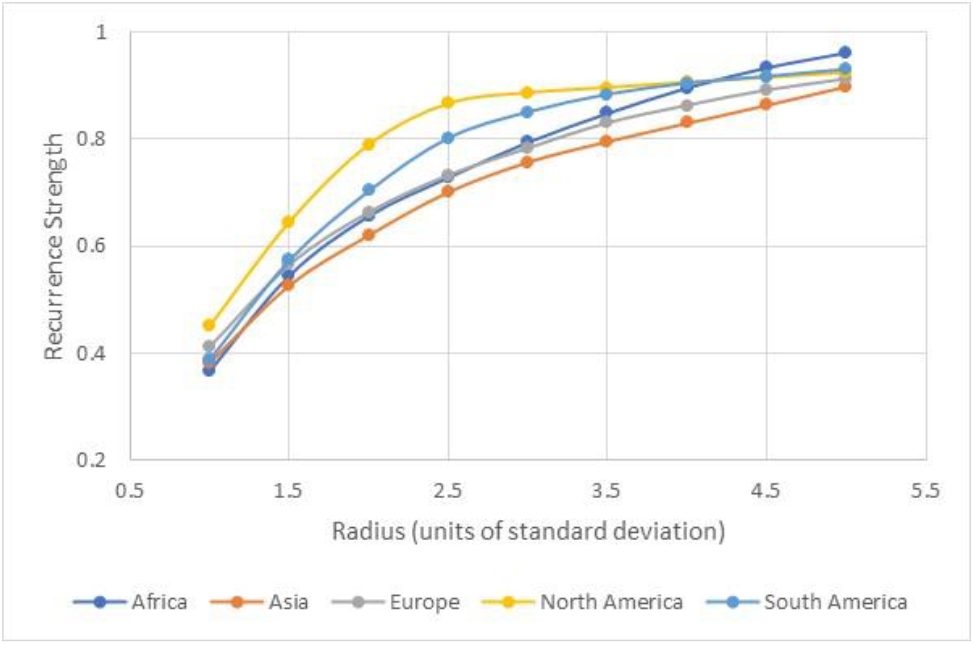
Average recurrence strength calculated for different radii with increasing radius, for the embedded series of the number of new cases per million with the radii taken in units of standard deviation.

We find that, for the lower values of the radius, North America stands out with a higher recurrence strength which initially grows faster than the remaining regions with increasing radius, this means that there is a stronger recurrence visible in lower radius values. For radii between 1.5 and 4 s.d. North and South America are the predominant regions in terms of average recurrence strength, that is they have the highest proportion of recurrence points over the number of diagonals below the main diagonal. Africa, in turn, only surpasses these two regions for radii of 4.5 s.d. and 5 s.d., with Europe and Asia converging from below to the North and South America’s average recurrence strengths as the radius is increased, but Asia stands out as being predominantly below the other regions for the analyzed radii.

The conditional 100% recurrence probability also shows a similar pattern. Asia stands out as the region with both the lowest average recurrence strength and the lowest probability of finding a 100% recurrence line in a random selection of lines with recurrence (figure 10). Contrastingly, North America dominates the intermediate radii, with Africa coming second up to the 3.5 s.d. radius, at a radius of 4 s.d., South America converges to North America’s pattern, and for radii of 4.5 s.d. and 5 s.d. Africa becomes dominant in terms of the probability of finding a 100% recurrence line conditional on the line containing recurrences points.

**Figure 10:**
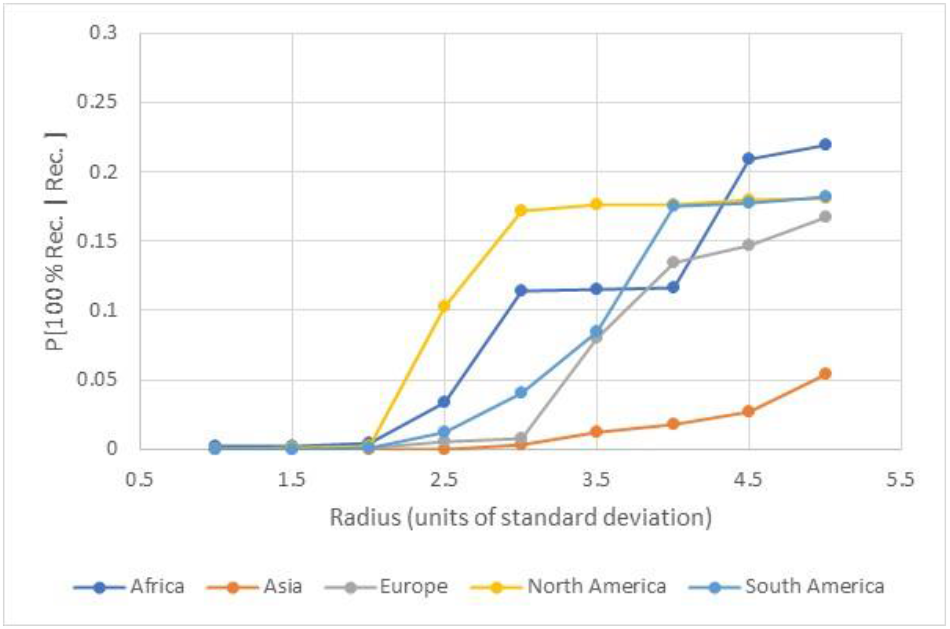
Conditional 100% recurrence probability calculated for different radii with increasing radius, for the embedded series of the number of new cases per million with the radii taken in units of standard deviation.

North America seems to have the strongest recurrence for intermediate radii, which implies that it may have the attractor that is closest to the *onset of chaos*, with the clearest recurrence structure. This pattern, however, is not the same for the new deaths per million’s reconstructed attractors as shown in figures 11 and 12 for the recurrence strength and conditional recurrence probability.

**Figure 11:**
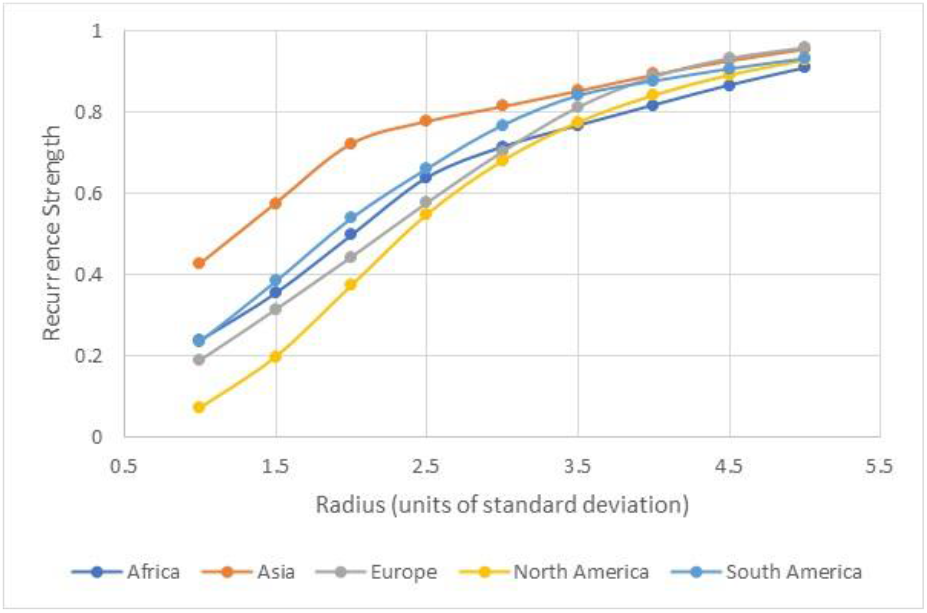
Average recurrence strength calculated for different radii with increasing radius, for the embedded series of the number of new deaths per million with the radii taken in units of standard deviation.

**Figure 12:**
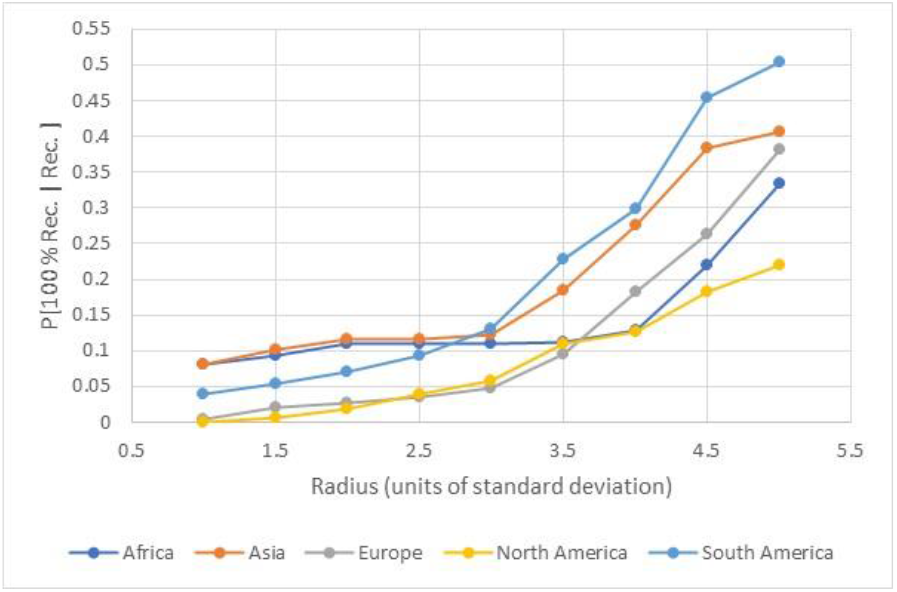
Conditional 100% recurrence probability calculated for different radii with increasing radius, for the embedded series of the number of new deaths per million with the radii taken in units of standard deviation.

Indeed, for the new deaths per million, we find that North America has predominantly lower recurrence strength and lower probabilities of finding 100% recurrence lines in lines with recurrence, this means that while North America’s new cases per million seems to have the strongest recurrence visible in intermediate radii, for the new deaths per million, we find that, until the radius of 3 s.d., Asia’s attractor is the one with the highest average recurrence strength and North America the one with the lowest, as the radius is increased beyond 3.5 s.d., Europe converges with Asia, while North America converges with South America, with Africa’s attractor becoming the attractor with the lowest value of recurrence strength though also convergent from below to the North and South America’s average recurrence strengths.

As for the probability of finding a line with 100% recurrence, conditional on the line being a line with recurrence, South America’s recurrence rises faster than the remaining regions, becoming the predominant line, which means that South America has a more regular structure when the radius is increased when it comes to deaths, followed by Asia, Europe, Africa and, finally, North America.

Considering, now, the homology classes analysis of the Vietoris–Rips filtration from the distance matrix *S*, we find that, in each region, the dominant dimension in terms of number classes is H0, which corresponds to connected components, followed by H1, which corresponds to loops and, finally, H2, which corresponds to voids (table 3). For the homology dimensions 1 and 2 we find that North America is the region with the highest number of classes, for the new cases per million.

**Table 3:** Number of persistence classes for the five regions and homology dimensions 0 to 2.

| Regions | New Cases |  |  | New Deaths |  |  |
| --- | --- | --- | --- | --- | --- | --- |
|  | H0 | H1 | H2 | H0 | H1 | H2 |
| **Africa** | 877 | 316 | 33 | 847 | 479 | 131 |
| **Asia** | 899 | 257 | 26 | 899 | 274 | 37 |
| **Europe** | 898 | 315 | 32 | 883 | 409 | 71 |
| **North America** | 899 | 337 | 44 | 869 | 458 | 164 |
| **South America** | 868 | 308 | 33 | 853 | 411 | 81 |

Indeed, for the number of new cases per million and the homology dimension 0, North America has the same number of classes as Asia (899), however, for the homology dimension 1, we find that North America has 337 classes while Asia only has 257 (which is the region with the lowest number of classes H1), a similar pattern is found for the homology dimension 2, where we find that North America has, again, the highest number of classes 44 while Asia only has 26, the lowest number of classes. This indicates that the North America’s new cases per million attractor may have a higher complexity in terms of its topological structure.

For the new deaths per million and H0, we find that Asia has the highest number of classes H0 (899) followed by Europe (883) with North America only coming in third with 869 classes. For H1, we find that Africa has the highest number of classes (479), followed by North America (458), while for H2 we find that North America has the highest number of classes (164) followed by Africa (131).

Considering the persistence metrics we find that, as shown in tables 4 and 5, for all regions, the new deaths per million show smaller values in both maximum and mean persistence than the new cases per million. For the new cases per million series, the lowest persistence values both in terms of maximum persistence and mean persistence are obtained for Africa and Asia, while Europe, North and South America show higher persistence metrics’ values for the homology classes. Regarding the maximum persistence values, we find that North America has the highest maximum persistence value of the group for homology dimension 0, but Europe has the highest values of terms of maximum persistence for homology dimensions 1 and 2, also standing out as having the highest mean persistence for all homology dimensions.

**Table 4:** Maximum persistence for the five regions and homology dimensions 0 to 2.

| Maximum Persistence | New Cases |  |  | New Deaths |  |  |
| --- | --- | --- | --- | --- | --- | --- |
|  | H0 | H1 | H2 | H0 | H1 | H2 |
| **Africa** | 12.7619 | 3.4790 | 0.9513 | 0.4419 | 0.1228 | 0.0253 |
| **Asia** | 46.8804 | 36.6568 | 2.8549 | 0.3554 | 0.2006 | 0.0305 |
| **Europe** | 447.1697 | **277.1158** | **32.5501** | 2.0519 | **1.2889** | 0.3488 |
| **North America** | **915.0522** | 183.8258 | 16.7437 | 3.7855 | 1.0945 | 0.3973 |
| **South America** | 248.9097 | 122.0910 | 8.0882 | **4.4615** | 1.2010 | **0.6799** |

**Table 5:** Mean persistence for the five regions and homology dimensions 0 to 2.

| Mean Persistence | New Cases |  |  | New Deaths |  |  |
| --- | --- | --- | --- | --- | --- | --- |
|  | H0 | H1 | H2 | H0 | H1 | H2 |
| **Africa** | 1.4334 | 0.3712 | 0.1373 | 0.0685 | 0.0106 | 0.0038 |
| **Asia** | 4.1626 | 1.3028 | 0.2843 | 0.0348 | 0.0084 | 0.0040 |
| **Europe** | **52.7557** | **14.2232** | **4.7414** | 0.5297 | 0.1263 | 0.0574 |
| **North America** | 40.0296 | 8.4910 | 3.3073 | **0.9573** | **0.1744** | **0.0717** |
| **South America** | 24.9506 | 6.3083 | 2.3986 | 0.6662 | 0.1367 | 0.0598 |

For the new deaths per million, we also find that Africa and Asia have the lowest persistence values both in terms of maximum persistence and mean persistence, again, with Europe, North and South America standing out. In regards to the maximum persistence metric and the homology dimension 0, we find that South America stands out followed by North America and by Europe, the same holding for homology dimension 2, for homology dimension 1, we find that Europe has the highest maximum persistence value followed by South America and then North America. Regarding the mean persistence, North America stands out as having the highest value.

For all groups there is only one infinity class which corresponds to homology dimension 0, furthermore, unlike the other dimensions, all homology dimension 0 classes are born at 0, as shown in figure 13 which shows the persistence diagrams for the new cases per million (top) and new deaths per million (bottom).

**Figure 13:**
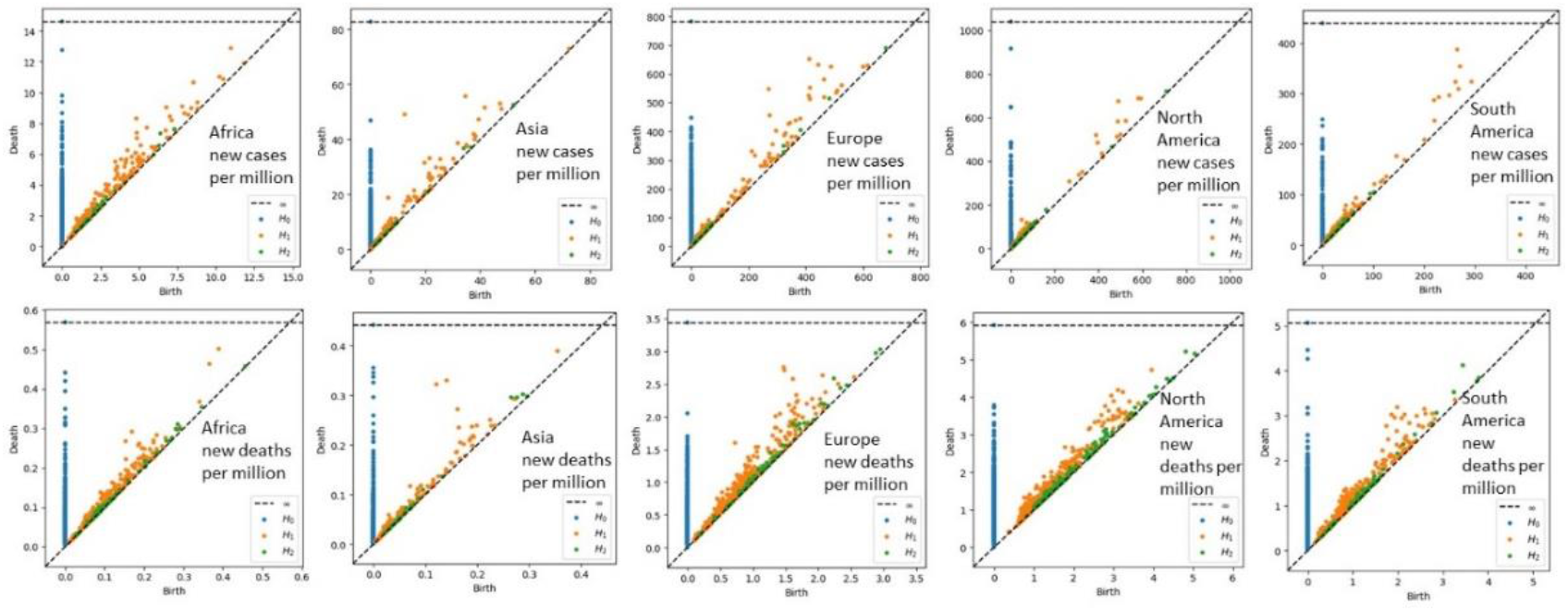
Persistence diagrams for each region’s number of new cases per million (top) and number of new deaths per million (bottom).

The persistence analysis, thus, shows some differences between the regions, in regards to the attractors’ topological structure, however, there is also a clear division into two groups: Africa and Asia, on the one hand, and Europe, North America and South America, on the other. The first group is characterized by smaller persistence metrics’ values, while the second is characterized by higher persistence metrics’ values. We also find that the new deaths per million show smaller persistence metrics’ values than the new cases per million.

Considering, now, the degree to which the recurrence patterns in a Euclidean distance matrix contain information that may allow an adaptive agent to use it to predict the corresponding series, as discussed in the previous section, we use the sliding window learning approach and scikit-learn’s radius neighbor learner. The key for the window choice is that it cannot be too short since we may not obtain any recurrence patterns, but it cannot also be too large since we will get a smoothed trajectory and the agent will not be able to predict well the performance.

In tables 6 and 7, we show the *R*^2^ score of the learner for a one-step-ahead learning using sliding learning windows between 7 data points (7 samples/one week) to 14 data points (14 samples/two weeks) for the new number of cases per million and the new number of deaths per million.

**Table 6:** *R*^2^ scores of the adaptive A.I. for a one-step-ahead learning of the number of new cases per million, for different sliding learning window sizes and each of the five regions.

| Learning Window | Africa | Asia | Europe | North America | South America |
| --- | --- | --- | --- | --- | --- |
| 7 | 82.01% | 91.68% | 88.87% | 77.31% | 82.38% |
| 8 | 80.92% | 90.87% | 87.84% | 74.47% | 80.76% |
| 9 | 79.05% | 89.95% | 86.15% | 72.87% | 78.32% |
| 10 | 77.47% | 88.98% | 84.82% | 71.27% | 76.05% |
| 11 | 76.78% | 88.03% | 84.16% | 69.15% | 74.57% |
| 12 | 76.77% | 87.24% | 84.41% | 67.37% | 74.13% |
| 13 | 76.90% | 86.48% | 84.98% | 68.11% | 74.07% |
| 14 | 76.46% | 85.54% | 84.85% | 66.17% | 73.17% |

**Table 7:** *R*^2^ scores of the adaptive A.I. for a one-step-ahead learning of the number of new deaths per million, for different sliding learning window sizes and each of the five regions.

| Learning Window | Africa | Asia | Europe | North America | South America |
| --- | --- | --- | --- | --- | --- |
| 7 | 82.02% | 91.72% | 84.17% | 62.84% | 88.51% |
| 8 | 81.68% | 91.42% | 82.62% | 60.27% | 87.76% |
| 9 | 80.63% | 91.00% | 80.44% | 55.65% | 86.49% |
| 10 | 79.69% | 90.54% | 78.70% | 51.97% | 85.44% |
| 11 | 79.12% | 90.04% | 77.90% | 51.38% | 85.09% |
| 12 | 79.09% | 89.51% | 78.71% | 54.32% | 85.72% |
| 13 | 79.28% | 88.99% | 80.24% | 58.83% | 86.76% |
| 14 | 79.06% | 88.38% | 80.50% | 60.45% | 87.05% |

The learning algorithm used is the radius nearest neighbor regression, which uses the recurrence information for a given radius to learn, in this way it allows us to see the predictability of the signal using the recurrence information from the reconstructed attractor, as discussed in the previous section. For the radius value we used five standard deviations (5 s.d.), the metric is the Euclidean distance and a brute force algorithm was used. The choice of the standard deviation comes out of the previous recurrence analysis, indeed for 5 s.d. we found the stronger recurrence results for all the regions, and the objective of this analysis is to characterize the degree to which the recurrence structure contains information on the future trajectory of each target series.

As can be seen in tables 6 and 7, for all the learning window sizes, the score is high showing that the past recurrence structure has a high degree of information on the dynamics for the one-period-ahead value of the corresponding target series.

However, the *R*^2^ score tends to decrease in general with the window, with the highest values of *R*^2^ being obtained for a 7 samples training window. In this case, North America stands out as the region with lowest *R*^2^ followed by Africa, which comes in second after North America as the region with the worst performance, even though the performance is very high for all the regions when a 7 period window is used, with all regions, except North America, exceeding an 80% *R*^2^, and with North America exhibiting an *R*^2^ of around 77.31% for the new cases per million and 62.84% for the new deaths per million. Noticeably, Asia stands out as having an *R*^2^ and explained variance higher than 90%, for both the new cases per million and the new deaths per million. Working, then, with a 7 samples window, we show in tables 8 and 9 the different metrics for the one-period-ahead prediction.

**Table 8:** Main performance metrics of the adaptive A.I. for a one-step-ahead prediction of the number of new cases per million, for a 7 samples sliding window.

|  | **Correlation** | **RMSE/Amplitude** | **Explained Variance** | **$R^2$ score** |
| --- | --- | --- | --- | --- |
| **Africa** | 0.905993 | 8.399% | 82.011% | 82.011% |
| **Asia** | 0.957695 | 4.989% | 91.709% | 91.680% |
| **Europe** | 0.942834 | 5.702% | 88.871% | 88.870% |
| **North America** | 0.879779 | 4.985% | 77.313% | 77.310% |
| **South America** | 0.908240 | 6.156% | 82.378% | 82.375% |

**Table 9:** Main performance metrics of the adaptive A.I. for a one-step-ahead prediction of the number of new deaths per million, for a 7 samples sliding window.

|  | **Correlation** | **RMSE/Amplitude** | **Explained Variance** | **$R^2$ score** |
| --- | --- | --- | --- | --- |
| **Africa** | 0.905970 | 7.175% | 82.019% | 82.019% |
| **Asia** | 0.957866 | 5.820% | 91.716% | 91.715% |
| **Europe** | 0.917579 | 8.784% | 84.174% | 84.174% |
| **North America** | 0.793074 | 10.769% | 62.837% | 62.837% |
| **South America** | 0.940868 | 7.281% | 88.515% | 88.515% |

Considering the correlation, we find a high positive correlation between the predictions and the series’ values for all the regions. This is indicative that the topological information contained in the 7 samples sliding window can be exploited by a radius neighbor algorithm to yield a forward-looking prediction for the one-period-ahead target variable with a very high performance in terms of linear correlation between the prediction and the target.

Again, North America stands out as the region where the artificial agent has the lowest performance, with the lowest correlation, albeit being close 0.88 in the new cases per million series and to 0.79 in the new deaths per million series. In regards to the RMSE/Amplitude in percentage indicator, we find that, for the one-step-ahead prediction, for the new cases per million, the highest error per amplitude is obtained for Africa with an RMSE that is around 8.399% of the total data amplitude, which is a low value, North America, in this case, shows the best performance of the group with a value of 4.985%, by contrast, for the new deaths per million series we find the worst performance in North America with a value of 10.769%, which means that the RMSE is around 10.769% of the total data amplitude, which is still a low result.

The explained variance and *R*^2^ score show a similar profile, with the best performance being obtained for Asia with a more than 90% value in these two indicators, for the two series, and the worst performance being obtained for North America with a near 77% value for the new cases per million and a near 63% value for the new deaths per million.

In terms of general pattern, we, thus, find that the one-period-ahead series has a strong predictability when using the topological information contained in the Euclidean neighborhood structure. Also, if we move beyond a single period prediction to consider a 1 week ahead (7 days), two weeks ahead (14 days) and a 30 days ahead prediction, the performance does not drop significantly as shown in tables 10 and 11, furthermore, it can even increase relative to the one-step-ahead prediction, as shown in table 11 for Europe, in which the performance increases for the 14 days ahead prediction and then again for the 30 days ahead prediction, for North America where there is an increase for the 14 days ahead prediction’s performance, with a subsequent decrease for the 30 days ahead prediction, and, finally, South America, for which there is a decrease in the 14 days ahead prediction’s performance and then an increase for the 30 days ahead prediction. The long range predictability of the series is to be expected given the black chaos spectrum and the high frequency periodicities.

**Table 10:**
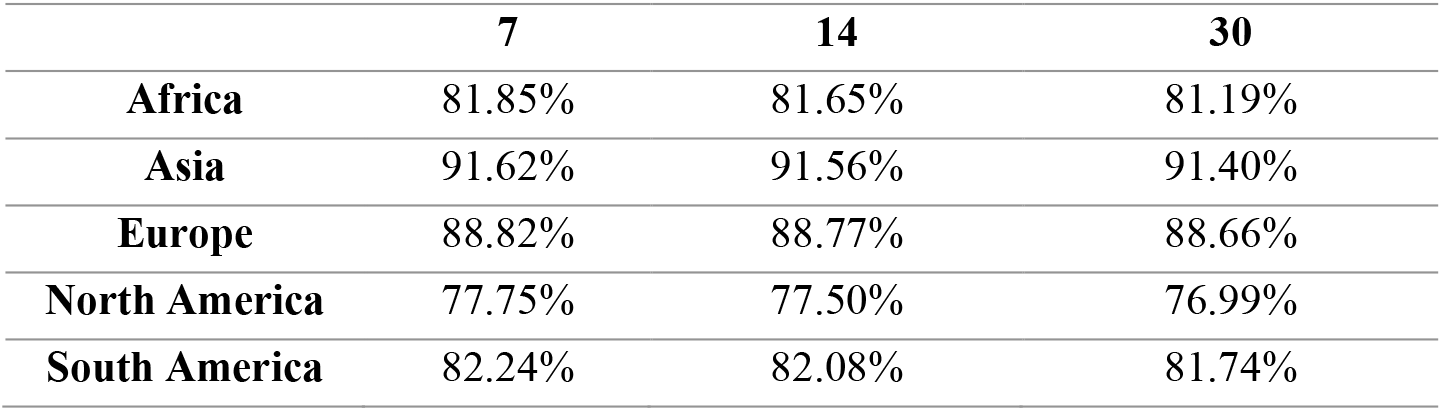
*R*^2^ scores of the adaptive A.I. for a 7, 14 and 30 days ahead prediction of the number of new cases per million, for a 7 samples sliding learning window.

|  | **7** | **14** | **30** |
| --- | --- | --- | --- |
| **Africa** | 81.85% | 81.65% | 81.19% |
| **Asia** | 91.62% | 91.56% | 91.40% |
| **Europe** | 88.82% | 88.77% | 88.66% |
| **North America** | 77.75% | 77.50% | 76.99% |
| **South America** | 82.24% | 82.08% | 81.74% |

**Table 11:** *R*^2^ scores of the adaptive A.I. for a 7, 14 and 30 days ahead prediction of the number of new cases per million, for a 7 samples sliding learning window.

|  | **7** | **14** | **30** |
| --- | --- | --- | --- |
| **Africa** | 81.89% | 81.74% | 81.44% |
| **Asia** | 91.64% | 91.55% | 91.35% |
| **Europe** | 84.21% | 84.51% | 84.79% |
| **North America** | 63.02% | 63.24% | 63.13% |
| **South America** | 88.45% | 88.44% | 88.48% |

Moving from the recurrence structure associated with the distance matrix to the *k*-nearest neighbors’ topological analysis, to perform this analysis, we first evaluate for which number of *k*-nearest neighbors the reconstructed attractor contains the most information that can be used to predict the series, in tables 12 and 13 we show the *R*^2^ scores for the single period prediction for the new cases per million and the new deaths per million. As can be seen from the results shown in both tables for Africa and Asia, the highest *R*^2^ is obtained only for a large number of nearest neighbors when compared to the sliding window size, while the highest score for Europe is achieved with a lower number of nearest neighbors.

**Table 12:**
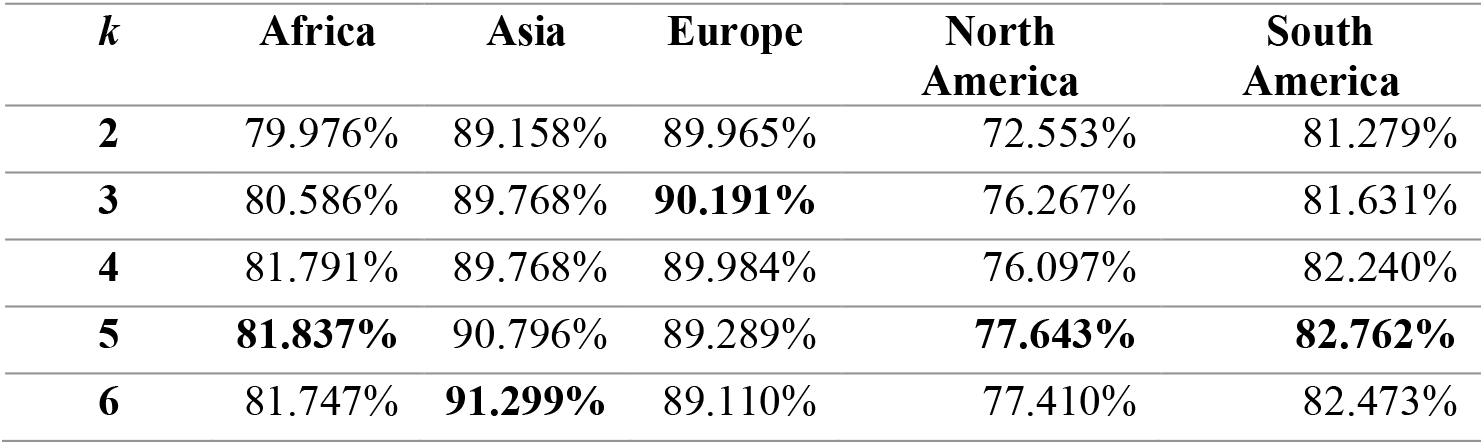
*R*^2^ scores of the *k*-NN adaptive A.I. for the new cases per million series and a 7 samples sliding learning window.

| **$k$** | **Africa** | **Asia** | **Europe** | **North America** | **South America** |
| --- | --- | --- | --- | --- | --- |
| **2** | 79.976% | 89.158% | 89.965% | 72.553% | 81.279% |
| **3** | 80.586% | 89.768% | **90.191%** | 76.267% | 81.631% |
| **4** | 81.791% | 89.768% | 89.984% | 76.097% | 82.240% |
| **5** | **81.837%** | 90.796% | 89.289% | **77.643%** | **82.762%** |
| **6** | 81.747% | **91.299%** | 89.110% | 77.410% | 82.473% |

**Table 13:** *R*^2^ scores of the *k*-NN adaptive A.I. for the new deaths per million series and a 7 samples sliding learning window.

| $k$ | Africa | Asia | Europe | North America | South America |
| --- | --- | --- | --- | --- | --- |
| 2 | 77.756% | 87.060% | **88.418%** | **75.759%** | 91.270% |
| 3 | 79.690% | 88.668% | 87.844% | 74.802% | **91.279%** |
| 4 | 80.426% | 89.768% | 87.147% | 70.697% | 90.489% |
| 5 | 81.009% | 90.569% | 86.203% | 67.331% | 89.726% |
| 6 | **81.596%** | **91.198%** | 85.012% | 65.443% | 89.043% |

For North America and South America the highest scores for the new cases per million are achieved for a higher number of nearest neighbors, while for the new deaths per million they are obtained for a smaller number of nearest neighbors. Globally, considering both series and all the regions, we find that the scores are very high, which means that the *k*-nearest neighbors also contain relevant information that can be used for prediction. Considering that the scores reflect the level of information that can be used for predicting the target series by an adaptive A.I. using the *k*-nearest neighbors’ algorithm, we should use, for the topological analysis, in each case, the number of neighbors that lead to the highest score in prediction, as discussed in the previous section, therefore, in figures 14 and 15 we show the *k*-nearest neighbor graphs and K-S entropies for each region, for the number of new cases per million (figure 14) and for the number new of deaths per million (figure 15), using, in each case, the number of nearest neighbors *k* that leads to the maximum *R*^2^ score shown in tables 12 and 13.

**Figure 14:**
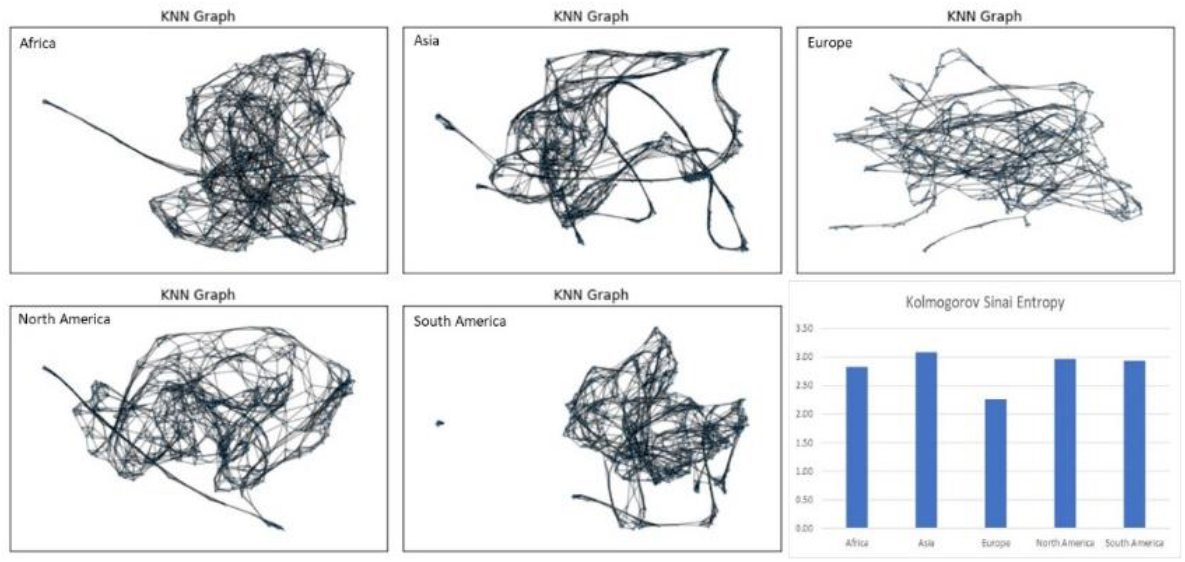
*k*-nearest neighbor graphs for each region’s number of new cases per million’s reconstructed attractor and the corresponding K-S entropy, using the value *k* that leads the highest *R*^2^ in table 12.

**Figure 15:**
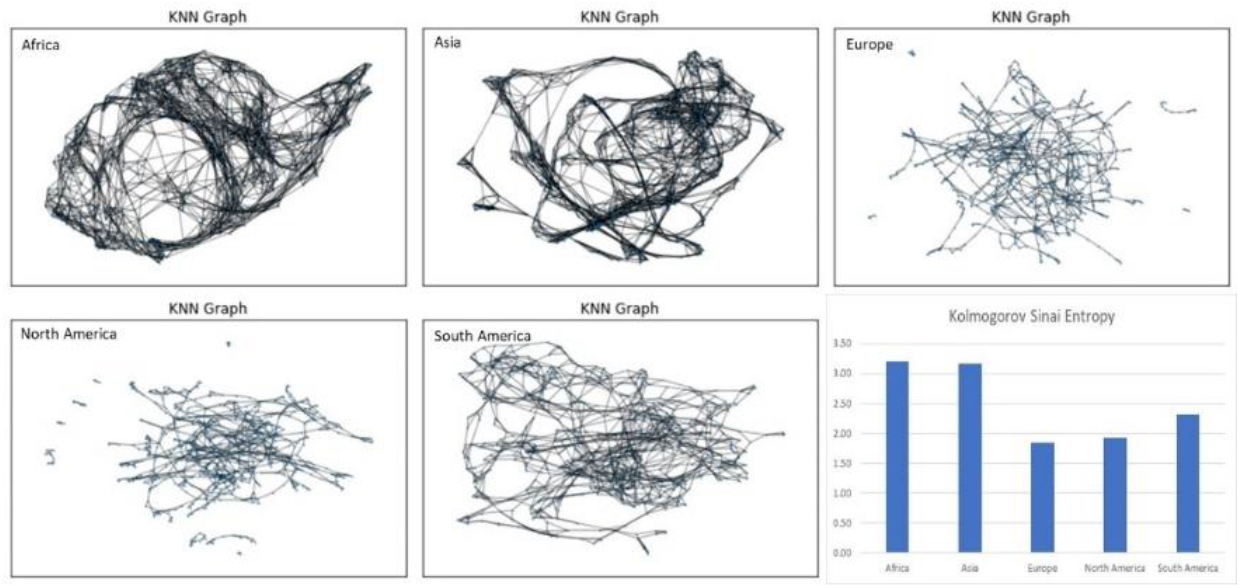
*k*-nearest neighbor graphs for each region’s number of new deaths per million’s reconstructed attractor and the corresponding K-S entropy, using the value *k* that leads the highest *R*^2^ in table 13.

As can be seen in figures 14 and 15, in conjunction with tables 14 and 15, we find that, for the new cases per million, in regards to the degree distribution entropy, all the regions exhibit a low entropy distribution with similar values (all close to 0.23), with the exception of Europe that stands out as the region with the lowest degree distribution entropy (around 0.19).

**Table 14:** Main entropy values for figure 14’s *k-*NN networks.

|  | Degree Distribution Entropy | Kolmogorov Sinai Entropy |
| --- | --- | --- |
| **Africa** | 0.231850 | 2.829805 |
| **Asia** | 0.237356 | 3.077790 |
| **Europe** | 0.191127 | 2.261513 |
| **North America** | 0.232304 | 2.960116 |
| **South America** | 0.231839 | 2.937294 |

**Table 15:**
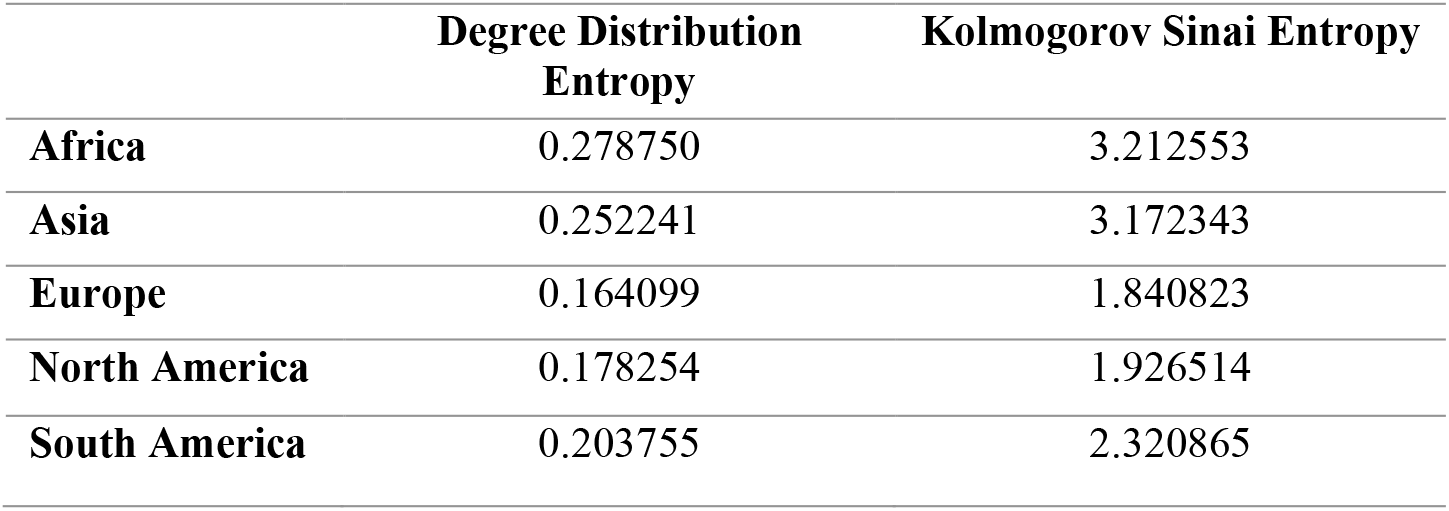
Main entropy values for figure 15’s *k-*NN networks.

Likewise, for the K-S entropy, all the regions show a similar profile (a value close to 3 bits) with Europe standing out with the lowest K-S entropy (around 2.26 bits). For the new deaths per million, we find again Europe as having the lowest entropy values but closer to those of North America.

Considering now the symbolic dynamics, we find that the ordinal partition graph for all regions, in the case of the reconstructed attractor, for the number of new cases per million, is the complete graph with six nodes *K*_6_ as shown in figure 16.

**Figure 16:**
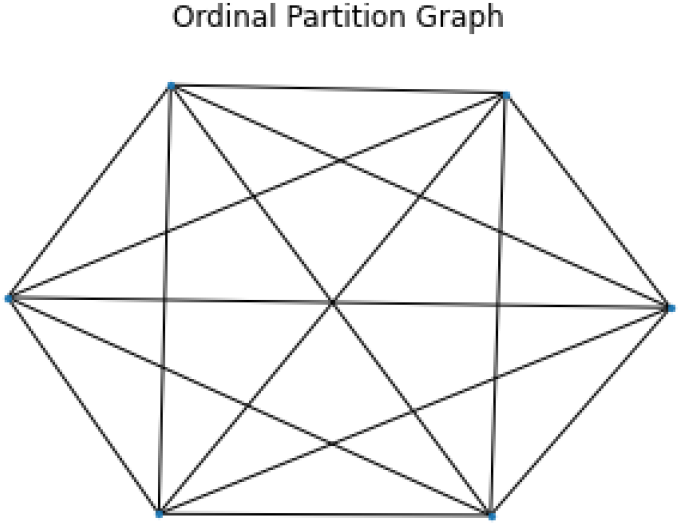
Ordinal partition graph that characterizes each region’s number of new cases per million’s reconstructed attractors.

The graph *K*_6_’s appearance in each region’s reconstructed attractor indicates that this is a robust topological feature of the SARS-CoV-2’s dynamics which is independent from each region’s dynamics. From a computational standpoint, such a *K*_6_ graph can be associated with a triadic encoding for a three-letter word with no repetitions, with each node associated with the possible permutations of a three letter word with each letter being chosen from a three letter alphabet, which provides for a symbolic encoding of the three dimensions.

The *K*_6_ structure arises due to two factors, the first is that the embedding dimension is equal to 3 which leads to six permutations, therefore six nodes for the graph, the second factor is that the dynamics is such that all possible transitions between the six nodes occur, which makes the graph for the transitions a complete graph.

From a symbolic dynamics standpoint, the fact that we get the same ordinal partition graph, means that we get a graph isomorphism between any two regions, all the regions’ reconstructed attractors are, thus, characterized by the same ordinal partition graph, and thus the same underlying computational structure, the permutation entropy values are also close to each other, however, there are some differences as can be seen in table 16.

**Table 16:** Permutation entropy for each region’s number of new cases per million’s reconstructed attractors.

|  | Perm. Entropy |
| --- | --- |
| **Africa** | 2.291354 |
| **Asia** | 2.209633 |
| **Europe** | 2.270652 |
| **North America** | 2.349743 |
| **South America** | 2.412355 |

As shown in table 16, Asia is the region with the smallest value for the permutation entropy, followed, in increasing order, by Europe and Africa. South America has the highest value of the permutation entropy followed by North America. The degree entropy for this graph is zero and the K-S entropy is the same for all regions and equal to 2.321928.

For the new deaths per million, the reconstructed attractor for Asia is still given by the *K*_6_ graph, with a higher permutation entropy 2.375796, without any change in the degree and K-S entropies’ values in comparison to the new cases per million attractor.

The remaining regions do not keep a complete graph structure for the new deaths per million, for their respective embedding dimensions, however, it should be stressed that, if a three dimensional embedding were used, then, we would still recover the complete graph structure for all regions, that is the *K*_6_ graph characterizes all regions at a three dimensional embedding. When the embedding dimension is risen beyond 3, we no longer get complete graphs.

In this case, for the embedding parameters obtained from the false nearest neighbors’ criterion, as shown in table 17, the region with the highest permutation entropy is North America followed by Africa, while in degree entropy we find Europe as the region with the highest entropy followed by South America, finally, with respect to the K-S entropy we find that South America is the region with the highest value followed by Europe. For better visualization, we show, in figure 17, the connected components of the *k*-nearest neighbors’ graphs for Africa, Europe, South America and North America.

**Table 17:** Ordinal partition graph main entropy measures for each region’s number of new deaths per million’s reconstructed attractors.

|  | Perm. Entropy | Degree Entropy | K-S Entropy |
| --- | --- | --- | --- |
| **Africa** | 5.784736 | 0.583711695 | 3.61576 |
| **Asia** | 2.375796 | 0 | 2.32193 |
| **Europe** | 3.678060 | 0.786350715 | 3.80668 |
| **North America** | 5.833933 | 0.581944959 | 3.74116 |
| **South America** | 3.966362 | 0.735372104 | 3.94223 |

**Figure 17:**
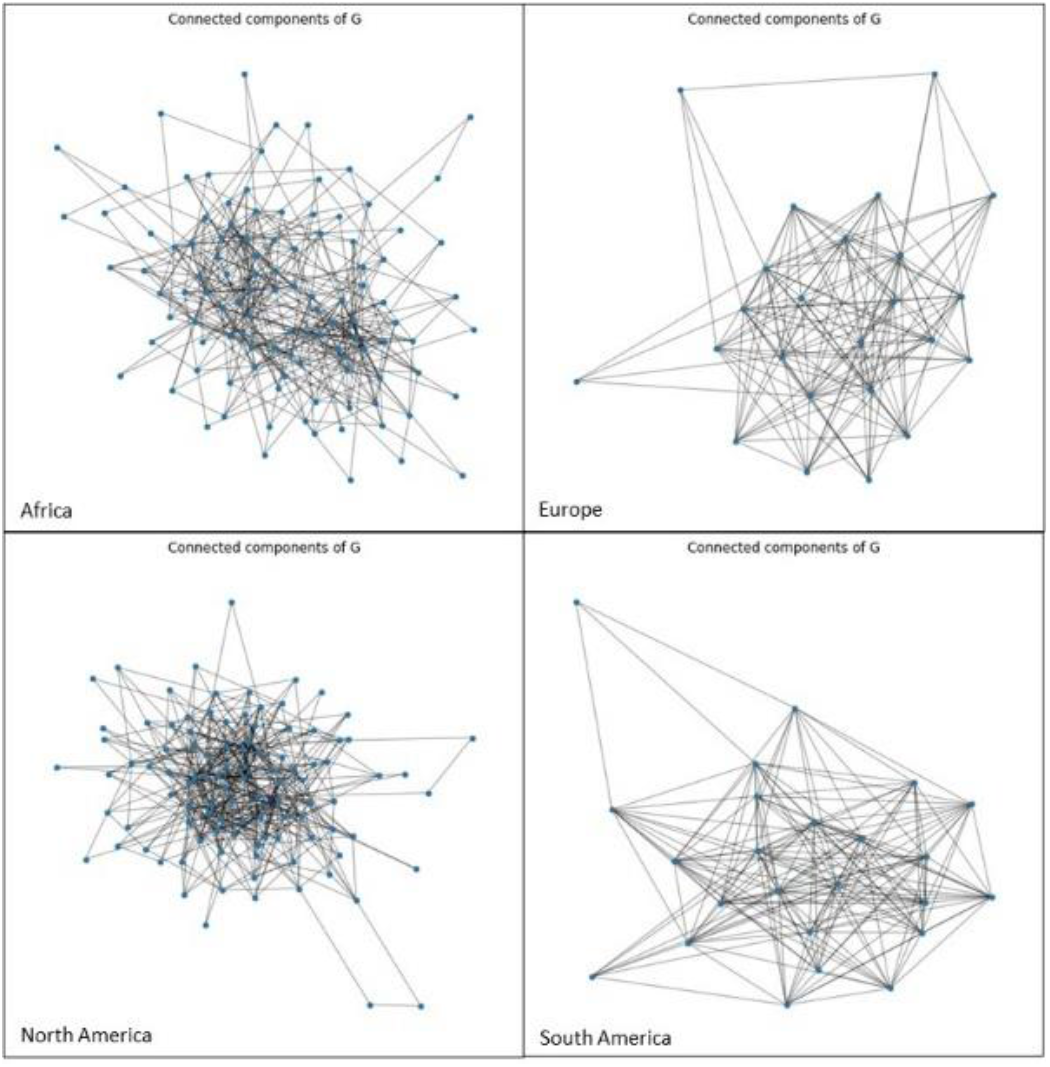
Connected components of the ordinal partition graphs that characterize Africa, Europe, North America and South America’s respective number of new deaths per million’s reconstructed attractors.

Having analyzed the cases where there is evidence of emergence of chaotic attractors, we now proceed to analyze the case of Oceania where there is evidence of a bifurcation.

### 3.2. Oceania’s Dynamics – a Bifurcation in SARS-CoV-2’s Outbreak Dynamics

Unlike the other regions, Oceania does not have a fixed attractor, there is instead evidence of a bifurcation associated with a break in an original attractor’s stability, followed rapid buildup of viral outbreak dynamics visible in both the new cases per million and the new deaths per million series.

The first marker of a change can be seen in the new cases per million and the new deaths per million time series charts as shown in figure 18, where two dynamics are clearly visible, first there is a lower fluctuation level, then there is a large outbreak followed by a turbulent dynamics with larger new cases per million values and a buildup in deaths.

**Figure 18:**
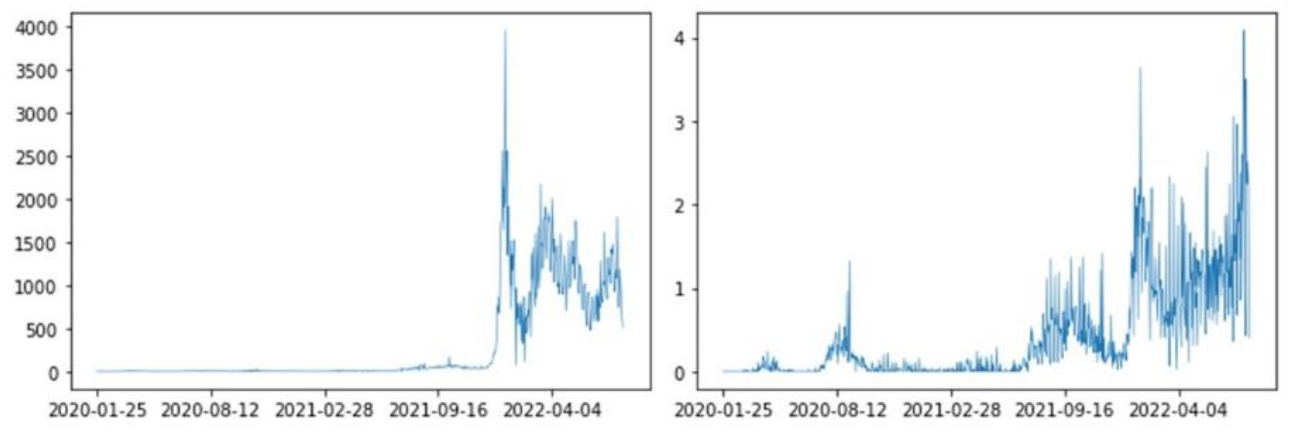
Oceania’s new cases per million series (left) and new deaths per million series (right).

The dynamics can actually be divided into three stages. The first stage is characterized by a dynamics with lower outbreak numbers, yet with markers of turbulence, this dynamics characterizes the period from 2020-01-25 to 2021-06-07. The second stage occurs between 2021-06-08 and 2021-12-14, which corresponds to a buildup stage leading to the large outbreak, in this period the initial dynamics and any corresponding possible attractor seem to have lost stability exhibiting a transient period with a buildup to the new more turbulent stage with higher number of reported positive cases and deaths, leading to the third stage, from 2021-12-15 onwards, where there is a large outbreak followed by an on average larger number of positive cases and deaths.

The three stages are shown in figure 19, for the number of new cases per million, which illustrates well the change occurring in the three stages.

**Figure 19:**
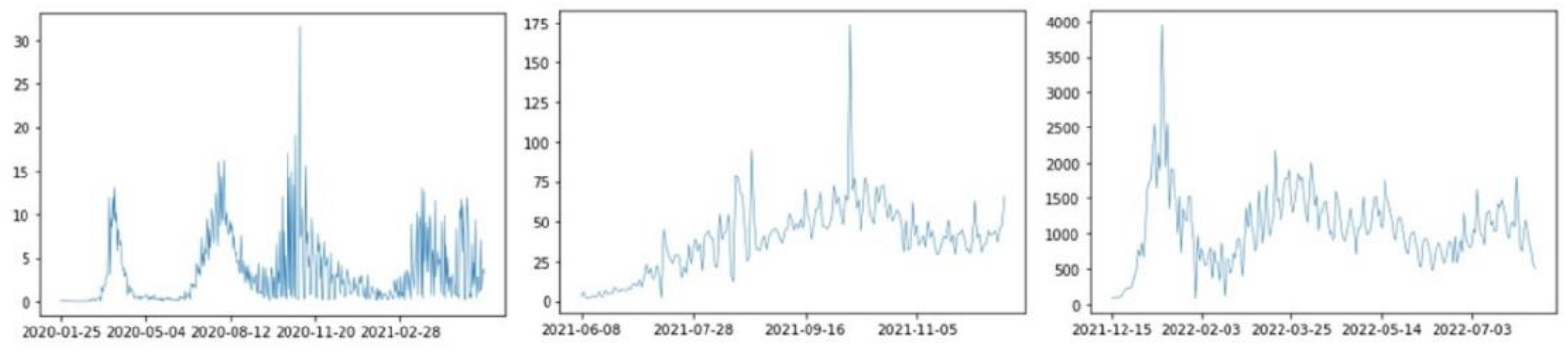
Three stages of the dynamics for the number of new cases per million, with stage 1 shown in the left, stage 2 shown in the middle and stage 3 shown in the right.

If we obtain the power spectral densities for each stage and the number of new cases per million, we find that stage 1 has a power law decay section in the low frequency range with a black noise spectrum with estimated exponent of 2.782218, with an associated *R*^2^ of 0.578568 and *p-value* of 0.000993, while stage 2 is characterized by the breakdown of the power law scaling with a fast transition to a white spectrum, stage 3 is again characterized by a black noise power law scaling in the spectrum with a power law decay with an estimated exponent of 2.960222, with an associated *R*^2^ of 0.717362 and *p-value* of 6.775415e-05. Figure 20 shows the power spectra for the three stages.

**Figure 20:**
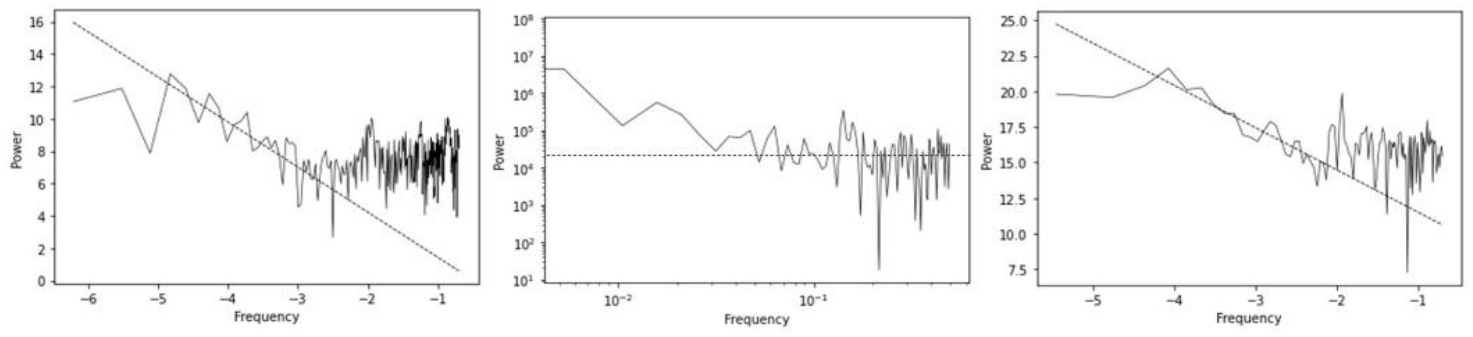
Power spectra for the three stages and the number of new cases per million, with stage 1 shown in the left, stage 2 shown in the middle and stage 3 shown in the right.

The evidence is compatible with a bifurcation occurring at the end of stage 1, with the loss of stability of the underlying dynamics. In stage 1, the dynamics is characterized by a black noise spectrum. In stage 2, there is a breakdown to a low memory process with a white spectrum as predominant, this marks a transient dynamics associated with a transition to a new dynamics, at stage 3, and a possible new attractor, which is characterized by a black noise spectrum with a breakdown to a white spectrum at the high frequency region. The new dynamics shows a stronger persistence than the previous one.

Faced with this evidence, we cannot use the false nearest neighbors’ selection process to obtain an embedding dimension, since there may be dimensional changes and the three stages do not provide for sufficient data for reliable embedding parameters and Lyapunov exponents estimations, a problem that is reinforced by the bifurcation itself which is contrary to the assumption of a fixed stable attractor assumed in these chaotic time series analysis methods. In this way, the chaotic time series metrics cannot be applied to the whole series. However, the topological analysis methods can.

Considering these methods, we find that there are some problems with using a recurrence matrix, in the sense that while it can be used to identify topological changes, the distances that may be used in a fluctuation scale change and this may interfere with the ability to extract recurrence statistics due to the identified bifurcation leading to different fluctuation sizes, in this case, it is best to use the *k*-nearest neighbors’ learner and compare the performance for different embedding dimensions, we still use the 15 day lag as in the other regions, taking into account the quarantine period effects. Considering, first, a base three-dimensional embedding with a 7 day sliding learning window, as shown in tables 18 and 19, we find that for the number of new cases per million the best performance is obtained for 6 neighbors, while, for the number of new deaths per million, the best performance is obtained for 5 neighbors.

**Table 18:**
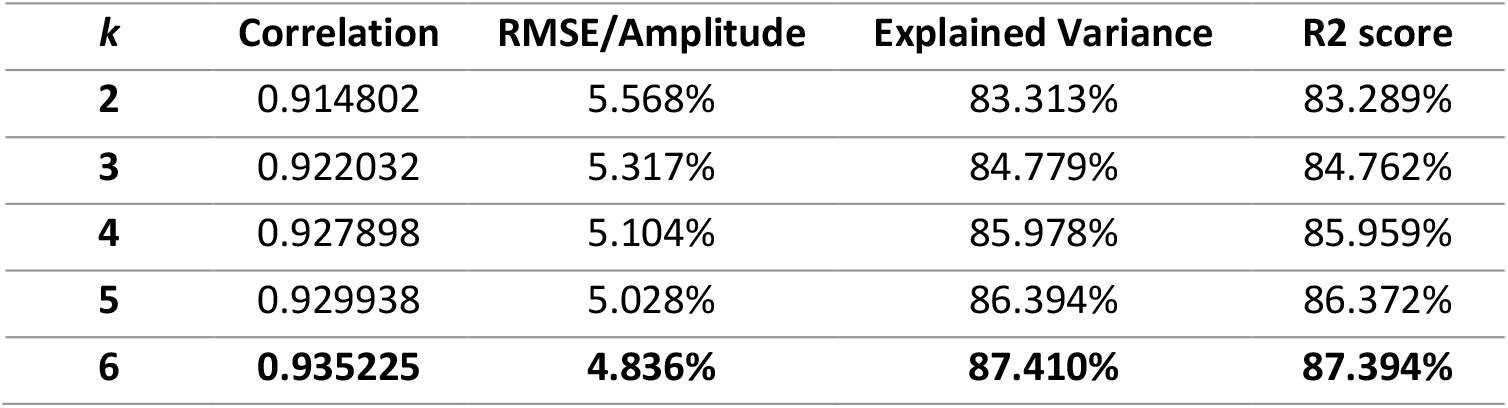
Performance scores, with a three dimensional embedding for the one period ahead prediction of a *k*-nearest neighbors algorithm for the number of new cases per million, with increasing values of *k* and a 7 sample sliding window.

**Table 19:** Performance scores, with a three dimensional embedding for the one period ahead prediction of a *k*-nearest neighbors algorithm for the number of new deaths per million, with increasing values of *k* and a 7 sample sliding window.

| $k$ | Correlation | RMSE/Amplitude | Explained Variance | R2 score |
| --- | --- | --- | --- | --- |
| 2 | 0.840435 | 8.116% | 70.496% | 70.300% |
| 3 | 0.828712 | 8.364% | 68.637% | 68.460% |
| 4 | 0.840793 | 8.082% | 70.678% | 70.551% |
| 5 | **0.845084** | **7.974%** | **71.363%** | **71.328%** |
| 6 | 0.843407 | 8.008% | 71.118% | 71.085% |

Taking these results into account, we compare the impact of increasing embedding dimensions on the predictability of the series, the dimension with the highest predictability is the one that we will use for further analysis, since it is the one that leads to the greatest captured pattern, including the bifurcation dynamics, which can be exploited by an adaptive A.I. using the *k*-nearest neighbors’ information, the chosen embedding dimension will be the one that has more information on the dynamics exploitable by such an agent.

As shown in tables 20 and 21, the best performance is obtained for a three-dimensional embedding. Using, thus, a three-dimensional embedding, we can apply the topological analysis methods for the *k*-nearest neighbors graphs and the ordinal partition graphs with the corresponding number of *k* nearest neighbors chosen from the values that lead to the highest prediction performance in tables 18 and 19, in this case, *k* = 6 for the number of new cases per million and *k* = 5 for the number of new deaths per million. In figures 21 and 22, we show, respectively, the corresponding *k* nearest neighbors and ordinal partition graphs for the number of new cases per million and the number of new deaths per million, for the three stages from left to right.

**Table 20:** Performance scores, for different embedding dimensions and one period ahead prediction of a *k*-nearest neighbors algorithm for the number of new cases per million, with increasing embedding dimensions and a 7 sample sliding window.

| dE | Correlation | RMSE/Amplitude | Explained Variance | R2 score |
| --- | --- | --- | --- | --- |
| **3** | **0.935225** | **4.836%** | **87.410%** | **87.394%** |
| **4** | 0.934814 | 4.881% | 87.324% | 87.310% |
| **5** | 0.935068 | 4.902% | 87.368% | 87.355% |
| **6** | 0.934272 | 4.962% | 87.212% | 87.199% |
| **7** | 0.933902 | 5.008% | 87.133% | 87.122% |
| **8** | 0.933633 | 5.048% | 87.081% | 87.070% |
| **9** | 0.933352 | 5.090% | 87.030% | 87.018% |
| **10** | 0.932736 | 5.145% | 86.911% | 86.899% |

**Table 21:** Performance scores, for different embedding dimensions and one period ahead prediction of a *k*-nearest neighbors algorithm for the number of new deaths per million, with increasing embedding dimensions and a 7 sample sliding window.

| dE | Correlation | RMSE/Amplitude | Explained Variance | R2 score |
| --- | --- | --- | --- | --- |
| **3** | **0.845084** | **7.974%** | **71.363%** | **71.328%** |
| **4** | 0.839036 | 8.155% | 70.360% | 70.292% |
| **5** | 0.836901 | 8.247% | 69.951% | 69.919% |
| **6** | 0.834142 | 8.361% | 69.469% | 69.441% |
| **7** | 0.834510 | 8.400% | 69.533% | 69.487% |
| **8** | 0.836365 | 8.394% | 69.865% | 69.820% |
| **9** | 0.834683 | 8.472% | 69.583% | 69.530% |
| **10** | 0.831927 | 8.578% | 69.098% | 69.040% |

**Figure 21:**
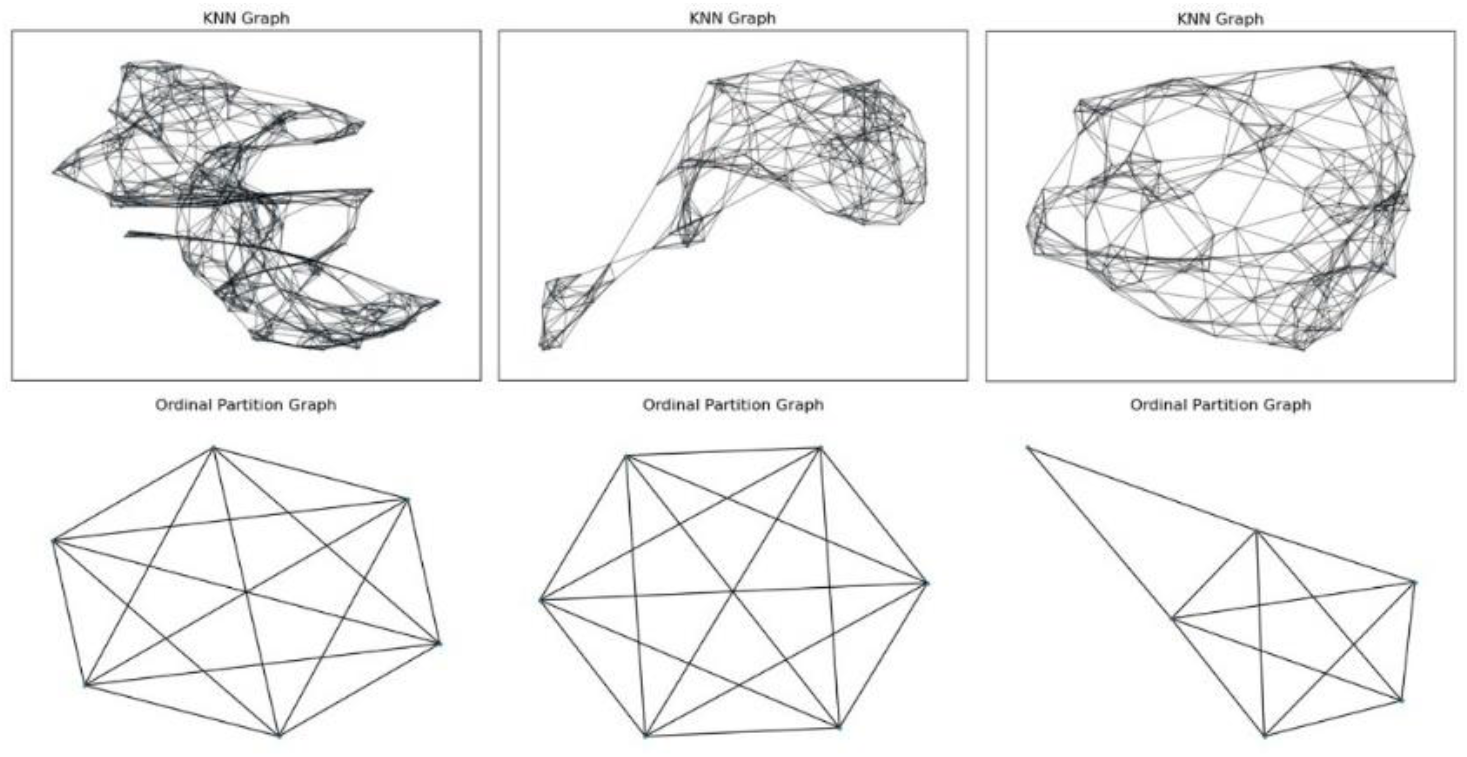
New cases per million’s *k* nearest neighbors (*k* = 6) and ordinal partition graphs obtained for a three dimensional embedding and with stage 1 shown in the left, stage 2 shown in the middle and stage 3 shown in the right.

**Figure 22:**
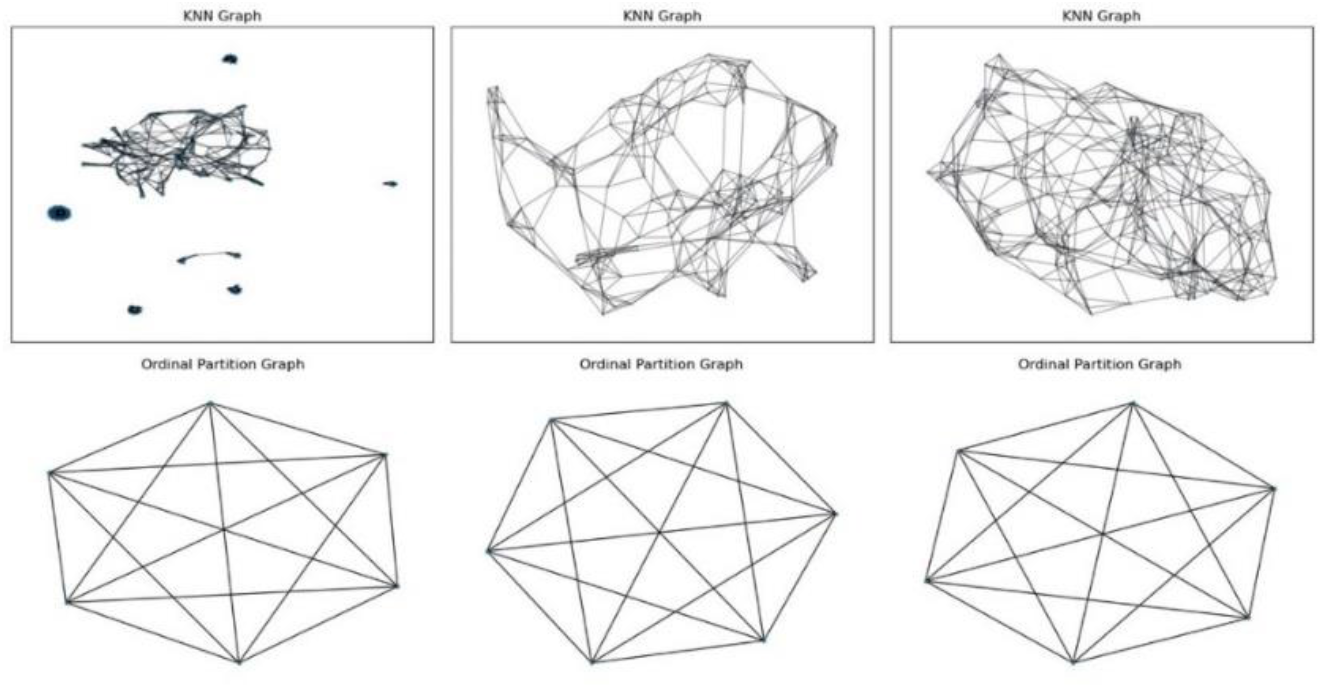
New deaths per million’s *k* nearest neighbors (*k* = 5) and ordinal partition graphs obtained for a three dimensional embedding and with stage 1 shown in the left, stage 2 shown in the middle and stage 3 shown in the right.

In tables 22 and 23 we show the respective graph entropy measures. Considering the new cases per million and the *k*-nearest neighbors graph, we find that the transition from stage 1 to stage 2 comes with an increase in the graph degree entropy but a decrease in the K-S entropy, which means that while there is a more random distribution of the node degrees, the entropy associated with a Markov process on the graph is reduced. From stage 2 to stage 3, still considering the *k*-nearest neighbors graph, we have a decrease in graph degree entropy and an increase in the K-S entropy. Therefore, while we have a less random distribution of the degrees, we have an increase in the entropy associated with a Markov process on the graph, which is the highest of the three stages.

**Table 22:** Entropy measures for the *k*-nearest neighbors and ordinal partition graphs of figure 21.

|  | Stage 1 | Stage 2 | Stage 3 |
| --- | --- | --- | --- |
| **Degree Entropy (KNN)** | 0.267476 | 0.330146 | 0.325345 |
| **K-S Entropy (KNN)** | 3.096103 | 3.077432 | 3.137238 |
| **Degree Entropy (Ordinal)** | 0 | 0 | 0.564475 |
| **K-S Entropy (Ordinal)** | 2.321928 | 2.321928 | 2.070895 |
| **Permutation Entropy** | 2.499085 | 2.408072 | 2.471680 |

**Table 23:** Entropy measures for the *k*-nearest neighbors and ordinal partition graphs of figure 22.

|  | Stage 1 | Stage 2 | Stage 3 |
| --- | --- | --- | --- |
| **Degree Entropy (KNN)** | 0.270277 | 0.294282 | 0.295310 |
| **K-S Entropy (KNN)** | 4.609673 | 2.762875 | 2.847433 |
| **Degree Entropy (Ordinal)** | 0 | 0 | 0 |
| **K-S Entropy (Ordinal)** | 2.321928 | 2.321928 | 2.321928 |
| **Permutation Entropy** | 2.228261 | 2.527406 | 2.556174 |

The ordinal partition graphs, in turn, provide important information, in stage 1 and stage 2 the ordinal partition graph is the *K*_6_ graph, which also characterizes the other regions’ new cases per million dynamics, as analyzed in the previous subsection. In this sense the Oceania dynamics at stages 1 and 2, from a topological standpoint in terms of the symbolic dynamics associated with the permutation analysis, shows a computational equivalence to the other regions, in the sense that we still get an ordinal partition graph that is isomorphic to the other regions.

Now, during stage 2, for the new cases per million, while we see the same *K*_6_ computational structure holding, there is a decrease in the permutation entropy, with respect to stage 1, finally, after the bifurcation, in stage 3, we no longer have the same type of topological structure as the other regions, from an ordinal partition standpoint, instead, while we still get a six node graph, as expected from the fact that we are performing a three dimensional embedding, however, only five nodes form a complete subgraph, this is the *K*_5_ graph, which corresponds to a pentacle, the sixth node is only linked to two vertices of the pentacle, forming a triangle, in this sense, the dynamics no longer performs every possible transition between all the permutations, three transitions are lost, with the number of links dropping from 15 to 12. This is unique in all the regions, since for all the other regions, whenever one performs a three dimensional embedding, the *K*_6_ graph is obtained.

Therefore, in stage 3, with respect to the new cases per million, we find a change in the number of connections, which leads to a rise in the degree entropy, since we no longer have a complete graph which has zero degree entropy, leading to a slightly greater diversity of connections, however, there is a decrease in the K-S entropy for the ordinal partition graph, there is also a rise in the permutation entropy with respect to stage 2.

For the new deaths per million, the loss of connectivity associated with the *K*_6_ graph does not occur, so that all of the stages are characterized by the *K*_6_ structure, in this sense the *K*_6_ computational framework is conserved, therefore, the degree entropy is zero throughout the three stages and the K-S entropy is also constant and equal to the *K*_6_’s K-S entropy. The permutation entropy, however, increases in each stage. Regarding the *k*-nearest neighbors, we find that there is also an increase in the degree entropy in each stage, however, the K-S entropy decreases in with respect to stage 1, showing lower values in stages 2 and 3, there is a slight increase in the K-S entropy in stage 3 but still with a lower value with respect to stage 1.

The bifurcation occurred with the outbreaks of different variants, including the delta in June and July 2021, which prompted lockdowns in Australia, and omicron variant which led to the large outbreak and is linked to the dynamics of the third stage [27,28].

More time needs to pass though for a more complete study of the dynamical markers of the third stage, including the possible sustainability of a possible new attractor or, alternatively, a possible sudden drop in cases linked to a new bifurcation or even a new bifurcation with a further rise in cases. The occurrence of bifurcations will largely depend on immunity versus the viral evolution with respect to the rise of new variants and subvariants and their respective infection and transmissibility properties.

## 4. Conclusions

We applied methods from chaos theory, topological data analysis and machine learning to the new SARS-CoV-2 cases per million and the new deaths per million series by region. For Africa, Asia, Europe, North America and South America, the evidence is favorable to the emergence of low dimensional noisy chaotic attractors for both series.

While there are some differences between the regions, they all exhibit evidence to the attractors being near the *onset of chaos*, with Africa, Asia and Europe also being characterized by power law decay in the power spectra characteristic of a form of color chaos, in this case black chaos, that is, chaos with a black noise-like spectrum. We found that there is enough information in the topological structure of these attractors for an artificial adaptive agent equipped with a nearest neighbor machine learning module to predict the future dynamics of each of these series using the reconstructed attractor, this holds for both radius and *k*-nearest neighbor modules.

For Oceania, we identified the occurrence of a bifurcation and characterized it using the artificial adaptive agent with a *k*-nearest neighbors’ learning module showing that the bifurcation can be well captured by the *k*-nearest neighbors’ sliding learning window, we used the highest predictability values for *k* to find the optimal parameters for topological data analysis and found that the bifurcation occurred with changes in *k*-nearest neighbors’ dynamics visible in the entropy metrics, but also, for the new cases per million, in the ordinal partition graph. This bifurcation was linked to new variants, namely delta and, in particular, the omicron variant.

The evidence that the attractors are near the *onset of chaos*, along with the occurrence of the bifurcation in Oceania that led to a large outbreak and a change in the dynamics to higher values of the number of new cases and new deaths per million, is indicative of the possibility that attractors may change due to bifurcations in the dynamics, particularly linked to the emergence of new variants with possibly larger transmissibility and with a lower immune response.

In this sense, from the results obtained from the analysis, we find that we do not have an homogeneous attractor for all the regions, different regions show topological differences, in terms of attractors and predictability, in the case of the persistent homology analysis, we found a division in two main groups: Asia and Africa on one side and Europe, North and South America on another side.

The evidence favorable to the hypothesis that attractors are near the onset of chaos reinforces the possibility of bifurcations, since, from the Oceania case, we know that bifurcations can happen and that they are linked to the emergence of new variants, this evidence implies that we do not have a dynamical self-organization to a long-term stable equilibrium associated with an endemic virus that has reached a stable infection profile, in this way, healthcare authorities will need to be attentive to new variants and possible containment measures as well as adjustments in vaccination policies and drug discovery for dealing with these new variants.

The current work, in what regards the Oceania region, shows that an adaptive agent with a sliding window learning is still robust in capturing dynamical changes in attractor stability and, conjointly with topological data analysis tools, may provide a way for healthcare authorities to monitor and anticipate possible bifurcations.

## Data Availability

All data produced are available online at https://ourworldindata.org/explorers/coronavirus-data-explorer

